# First clinical experience with a novel optical-ultrasound imaging device on various skin pathologies

**DOI:** 10.1101/2021.06.28.21259325

**Authors:** Gergely Csány, L. Hunor Gergely, Klára Szalai, Kende K. Lőrincz, Lilla Strobel, Domonkos Csabai, István Hegedüs, Péter Marosán-Vilimszky, Krisztián Füzesi, Miklós Sárdy, Miklós Gyöngy

## Abstract

**Objectives:** A compact handheld skin ultrasound imaging device has been developed that uses co-registered optical and ultrasound imaging to provide diagnostic information about the full skin depth and lesions encountered therein. The aim of the current work is to present the first clinical results of the device. Using additional photographic, dermoscopic and ultrasonic images as reference, the images from the device are assessed in terms of the detectability of the skin layer boundaries (between the epidermis, dermis, and subcutis), and in terms of image features produced by common skin lesions.

**Methods:** Combined optical-ultrasonic recordings of various types of common skin lesions (melanoma, basal cell carcinoma, keratosis, dermatofibroma, naevus, dermatitis, psoriasis) were taken with the device (N = 53) and compared with images from a reference portable skin ultrasound imager. The investigator and two additional independent experts evaluated and compared the images in terms of skin structure detectability and skin lesion features.

**Results:** Skin structure detectability was unanimously over 90 % for epidermis, dermis and lesion. Morphological and echogenicity information observed for melanoma, basal cell carcinoma, keratoses, dermatofibroma, naevi, atopic dermatitis, psoriasis were found consistent with those of the reference ultrasound device and relevant ultrasound images in the literature.

**Conclusions:** The presented device is able to obtain simultaneous in-vivo optical and ultrasound images of common skin lesions. This has the potential to provide relevant information in a number of settings to be investigated in the future, including preoperative planning of skin cancer treatment.

## 1. Introduction

### 1.1. Motivation

As a result of continuous technical developments, various non-invasive imaging procedures are becoming increasingly adopted in dermatology.^1^ Non-invasive imaging devices that also examine the layers of the skin in depth open up new possibilities, especially in the areas of early diagnosis, treatment planning, follow-up, and teledermatology. These techniques include ultrasound (US) imaging, which is playing an increasingly important role in dermatology.^2^ There is a general trend in ultrasound to make devices more portable and user-friendly, while maintaining image quality. The aim of the current work is to present preliminary clinical experience with such a novel device for skin ultrasonography, which in addition also records co-registered optical images of the skin surface.

Skin cancer is one of the most common cancers in developed countries. In the United States, it is the most common type of cancer, affecting 20 % of the population in their lifetime.^3^ Early and accurate screening, and appropriately chosen, planned and implemented treatments play a key role in preventing malignant skin cancer deaths.^4^ Inflammatory skin diseases such as atopic dermatitis and psoriasis are also common skin maladies. Atopic dermatitis affects roughly 10 % of the population in developed countries.^5,6^ 40 % of the patients affected have moderate to severe symptoms, which means that, on average, they spend up to a third of their time fighting inflammation.^7^ Ultrasound imaging has the potential to play an important role in improving the detection, treatment planning and follow-up of the above-mentioned skin diseases with significant prevalence.^8-13^ Combining the ultrasound information with optical imaging can further strengthen diagnostic accuracy,^14^ since ultrasound information alone may not be enough for the diagnosis of skin diseases such as skin cancer.^15^

### 1.2. Overview of skin ultrasound

Ultrasound imaging makes it possible to map and objectively examine the subsurface structures of human tissues in a safe and cost-effective manner, without the use of ionizing radiation. Compared to more established ultrasound applications such as abdominal imaging, skin ultrasound uses higher frequencies (>15 MHz), with the depth of penetration posing a limit to the frequency that can be used. At a frequency of around 20 MHz, a depth of 6–7 mm and a resolution of 50–200 μm can be achieved.^16^ The examination of structures under the surface of the skin provides useful information for dermatologists and plastic surgeons in many cases. These include tissue structures, morphology, spatial extent, and location of various lesions.^2^ High-frequency ultrasound imaging has the potential to improve the accuracy of the diagnosis of certain skin diseases (proven malignant skin lesions),^17-19^ as well as the planning of different interventions and the following of treatment results via quantitative examination of the depth extent and depth spreading of the lesions.^20,21^ Due to the above advantages and applications, the use of ultrasound in dermatology is emerging,^22^ but due to the cost of the above-mentioned high-frequency devices currently available and the difficulty of interpreting skin ultrasound images (need for radiological expertise), ultrasound skin examination is mostly only available in larger medical centers. Given the workload on the centers (centers may have month-long waiting times),^23^ it would be useful if dermal ultrasound technology were more widely available to dermatologists and plastic surgeons. It is essential for the above aim to reduce the cost, increase portability, and improve and extend the interpretability of the images for non-radiologist users of skin ultrasound imaging devices.^24^

### 1.3. Aim and overview of current work

A recent paper by Mlosek et al.^25^ provides a valuable review of current skin ultrasound devices, noting that dedicated skin ultrasound scanners have higher portability than classical ultrasound scanners, making them better-suited for skin examinations. The aim of developing the *Dermus SkinScanner* was to take this portability and usability further. The *Dermus SkinScanner* is currently a premarket device, with a novel feature of combining optical and ultrasound imaging in an integrated, handheld device. This feature aids precise positioning of the ultrasound recordings, which in turn potentially aids reproducibility of the examinations, an aim expressly stated in a recent position statement of the European Federation of Societies for Ultrasound in Medicine and Biology (EFSUMB) on dermatologic ultrasound (Position Statement 5).^22^

It is the first time that clinical images of this device are presented in the literature. The remaining sections are structured as follows. In Section 2, a brief overview of the device itself is provided, as well as details of the clinical study. Section 3 first presents the detectability of skin structures using the device, as compared with a reference device. The remainder of the section shows representative images of common skin lesions caused by tumours and inflammatory skin diseases. Section 4 provides a discussion of the results in the context of the literature, as well as the potential uses of optical-ultrasound imaging provided by the *Dermus SkinScanner* and needs for further development and research. In Section 5, appropriate conclusions are drawn regarding the suitability of the device to image skin structure and common skin lesions.

## 2. Materials and Methods

### 2.1. The Dermus SkinScanner device

The *Dermus SkinScanner* is a wireless, cost-effective, compact portable optical and ultrasound imaging device that is developed by Dermus Kft. (Budapest, Hungary). The compact device contains the imaging, data processing and displaying systems together with its own battery-based power supply. It is easy to handle, relatively light-weight (500 g) and provides the user interface (including the display of recordings) at the point of the examinations, making it suitable for widespread everyday use in dermatology. The *Dermus SkinScanner* also contains an optical imaging module that helps positioning and thus also reproducibility of ultrasound imaging.

The *Dermus SkinScanner* uses a single-element ultrasound transducer with 33 MHz nominal center frequency. Two-dimensional ultrasound imaging is realized via mechanical scanning applying a physical slider on the device and an automated scan conversion algorithm,^26^ verified using phantoms and dermatological data in previous work.^27^ The imaging window is covered by a silicone membrane. Ultrasound gel is applied on the membrane surface for scanning.

As mentioned above, a unique feature in comparison to other portable ultrasound imaging devices is that *Dermus SkinScanner* uses optical guidance for the enhancement of precise positioning and repeatability of the recordings. In practice, this means that real-time optical images of the area of scanning are displayed with 10-fold magnification during positioning, before scanning. The results of a single recording are then displayed in terms of a 2-D ultrasound image and the corresponding optical image of the surface, with a marker indicating the relative position of the ultrasound image slice to the surface. The ultrasound image is colorized with a colour scale that provides a stronger contrast than those of the conventional grayscale representation and is aimed to facilitate the interpretation of the clinical information for non-radiologist users. Both the optical and ultrasound images of the device are automatically and anonymously saved using a cloud system.

Figure 1 presents photos of the device, with its various components highlighted.

**Figure 1.**
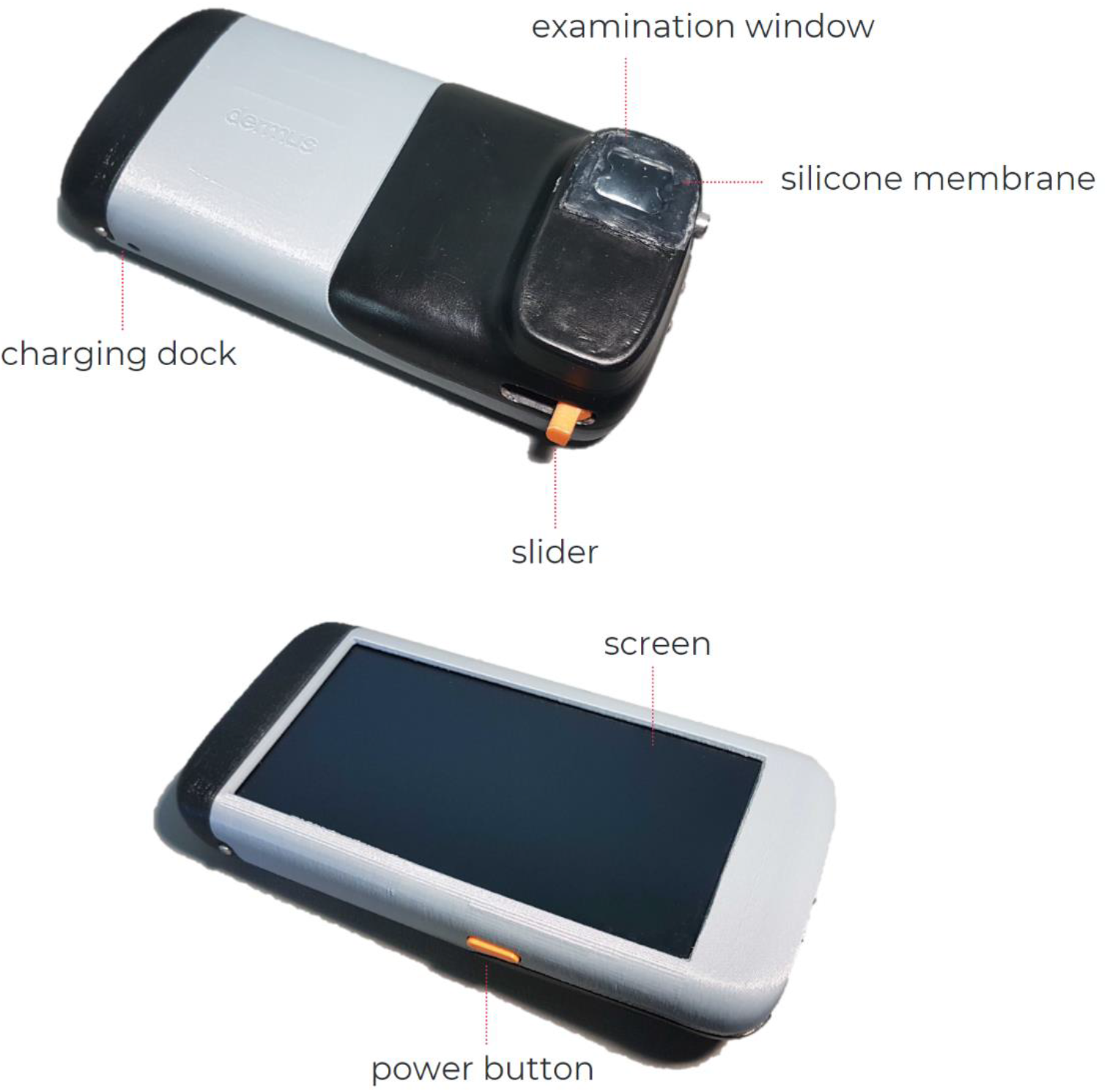
The Dermus SkinScanner device with its main components, including an examination window with a silicone membrane cover; screen; charging dock; power button; slider for mechanical scanning).

### 2.2. Study design

The present study was performed under the ethical approval of the Hungarian National Institute of Pharmacy and Nutrition (approval number OGYÉI/8317/2020), with all subjects providing their written informed consent. The examination site was the Department of Dermatology, Venereology and Dermatooncology, Semmelweis University, Budapest, Hungary. The examinations were performed by a dermatologist professional, hereinafter referred to as the “investigator”.

#### 2.2.1. Study population

The study included patients with a relatively wide range of skin pathologies from a representative population. The patients were primarily chosen from daily outpatient care, based on the criteria of the presence of skin lesions that were affecting the dermis and/or epidermis and that could be examined with ultrasound, and of being exempt from the exclusion criteria. Exclusions were limited to the presence of any signs of damaged, bleeding or purulent skin surfaces or skin lesions in the area to be examined, the location of the lesion being over an eyelid, withdrawn consent of the patient, or the presence of any disease that would endanger or interfere the examination in the opinion of the examining physician.

The patient population included both women (38 %) and men (62 %), with a quasi-uniform distribution of ages between 23 and 87 years for both genders.

In total, 53 lesions were examined of 39 patients. The vast majority (91 %) of the lesions were benign lesions or malignant skin tumours (including hemangiomas, naevi, keratoses, basal cell carcinomas and melanomas) and a smaller proportion (9 %) consisted of skin inflammatory lesions (related to dermatitis, psoriasis or morphea scleroderma). Regarding the locations of the lesions, the skin of several body regions have been examined, including those of the head, face, nose, upper and lower limbs, chest, abdomen and back.

#### 2.2.2. Data acquisition

Several optical and ultrasound images were taken of each lesion during the examinations. First, a clinical (macroscopic) photo and a dermatoscopic photo were taken of the skin lesion. This was followed by ultrasound recordings using two different portable skin imaging devices: the *Dermus SkinScanner* (as described in Section 2.1) and a reference device, the *Dramiński DermaMed* (briefly described below).

The *DermaMed* (*DRAMIŃSKI S*.*A*., Olsztyn, Poland) is a relatively light-weight (300 g), high-frequency (dual-mode 30/48 MHz, used in the 48 MHz mode) ultrasound imaging device that requires a USB-cable connection to a PC that is used for power supply and for the processing, display and storage of two-dimensional grayscale ultrasound images (or series of such images). The probe uses tilting mechanical scanning with a single-element transducer moved by an electrical motorized scanning system in a chamber that should be filled with distilled water and closed by a stretched film by the user, for appropriate coupling.

As described in Section 2.1, the optical image of the *Dermus SkinScanner* stored the information of the spatial position and orientation of its ultrasound scans in relation to the surface of the skin area of examination. This information was not available for the *Dramiński DermaMed* recordings, nevertheless they were taken with keeping the spatial orientation of its scans as close to those of the other device as possible. However, a perfect precise alignment was not possible due to the probe physically covering the area of examination during scanning.

To present the results of examinations of specific cases, the corresponding figures show the images (of various imaging modalities and devices, as described above) in a consistent layout throughout this article. The layout is presented below in Figure 2. The primary layers associated with the skin (epidermis, dermis, subcutis) as well as the lesions themselves are indicated by certain markers as an aid for the interpretation of the images throughout this article.

**Figure 2.**
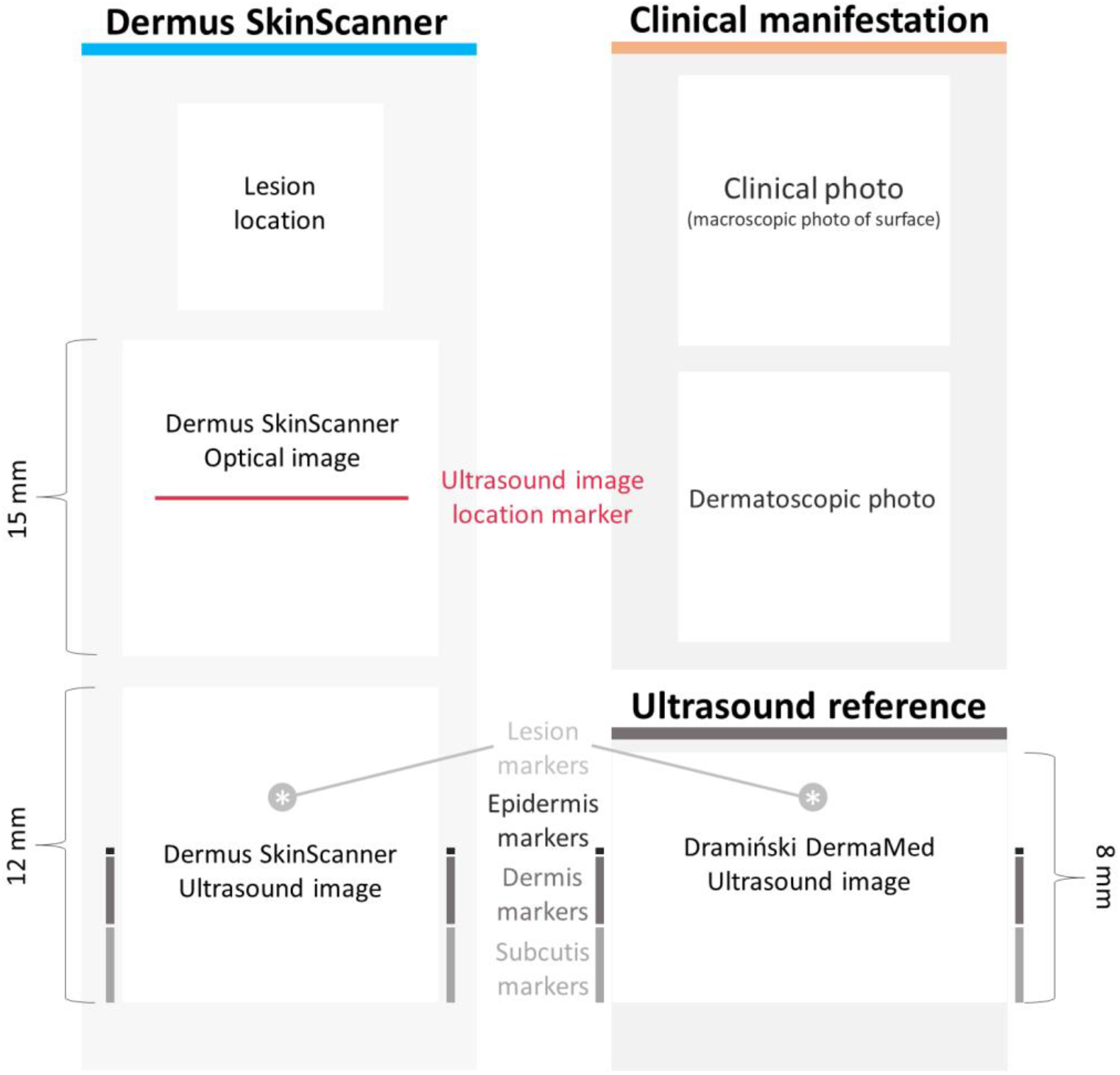
The layout in which the results of examinations are presented in this article. Image information provided by the Dermus SkinScanner device is presented on the left: lesion location (as marked by the investigator, on a body schematic), optical image with a red line marker representing the location of the corresponding ultrasound image slice, and the corresponding ultrasound image. On the top right, photo-documentation of the visual information seen by a dermatologist on a basic examination is presented: clinical (macroscopic) and dermatoscopic photo of the lesion. The corresponding ultrasound image of the reference device is presented on the bottom right in the layout. The primary skin layers and the lesion itself are marked on both ultrasound images as an aid for the interpretation of the images. Scaling of the images is also presented on this figure for reference (the aspect ratio is 1:1 for all of the images). The lesion markers (*) are placed just above the lesion.

**Figure 3.**
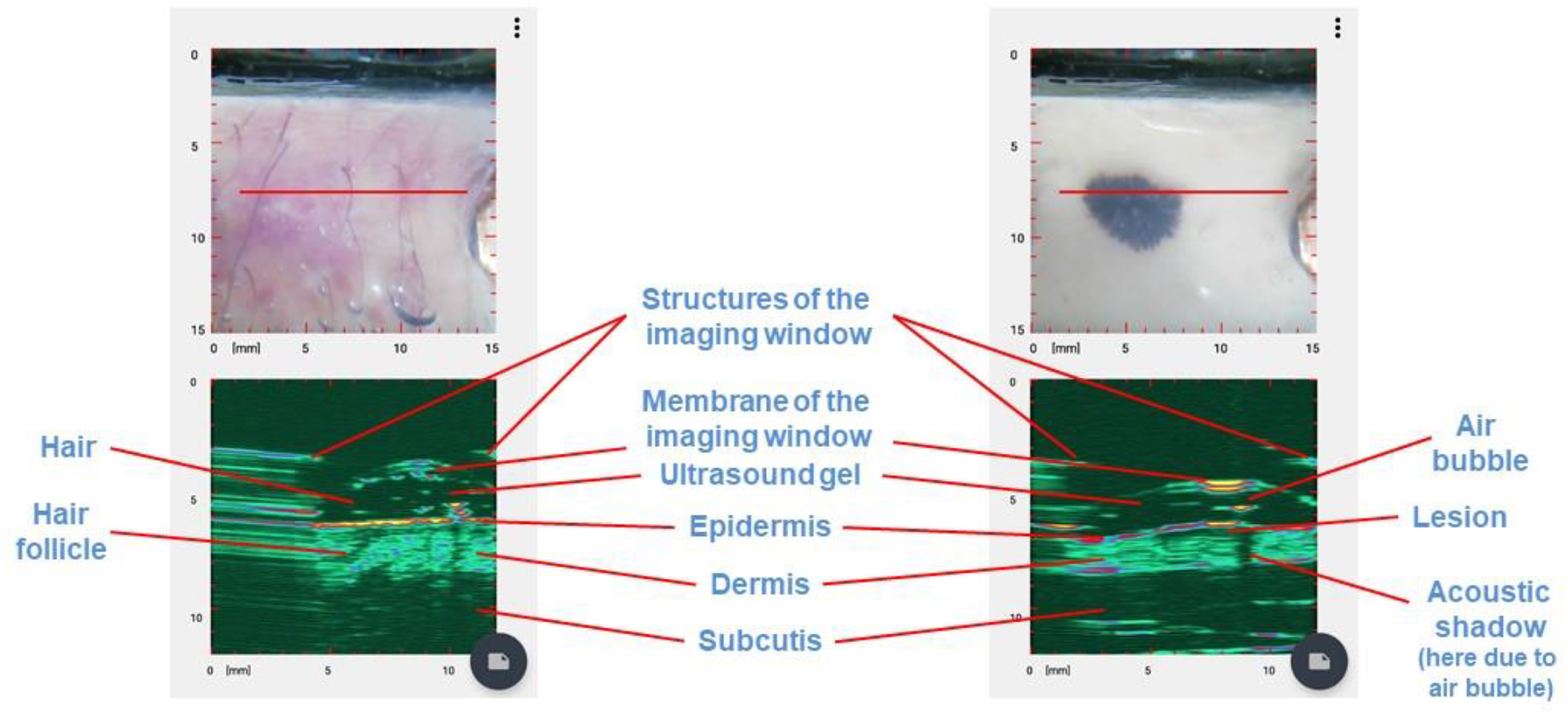
Typical elements of the ultrasound images of the Dermus SkinScanner. Ultrasound reflections from skin structures outline layers like the epidermis, dermis and subcutis, and also other structures like lesions or hair follicles. These structures are preceded by reflections from the membrane and other structures of the imaging window of the device itself. The ultrasound gel filling the volume in between the imaging window membrane and the surface of the skin is reflection-free; however, reflections from hair or air bubbles may be present in this region, and they may even cause the presence of acoustic shadows on the deeper regions of the image.

#### 2.2.3. Evaluation of skin structure detectability

The ultrasound images taken with the *Dermus SkinScanner* were evaluated and compared to the ones obtained with the *Dramiński DermaMed*, as a reference, in terms of the detectability of structural elements of the skin, such as the epidermis, dermis, subcutis and lesion. The evaluation was performed by both the investigator (on site) and by two independent experts (off site).

Since the use of the *Dermus SkinScanner* device aims to exploit the novel field at the intersection of dermatology and radiology, the two independent experts were chosen for the evaluation as representatives from these two medical specialties to cover the interdisciplinary field of dermatologic ultrasonography. The independent experts included a dermatologist with limited experience in skin ultrasound imaging, and a radiologist with extensive experience (10+ years) in skin ultrasound imaging. The two independent experts did not participate in any of the examinations and also did not have previous experience with the ultrasound devices used in the study.

## 3. Results

Following a summary of skin structure detectability and a brief section on interpreting the SkinScanner optical-ultrasound images, representative images of various common skin lesions are presented (using the layout described in Section 2.2.2). For the cases shown in Figures 4, 5, 7, 8, 11, 16, histopathological diagnosis was also available, which always confirmed the preliminary diagnosis of the investigator.

**Figure 4.**
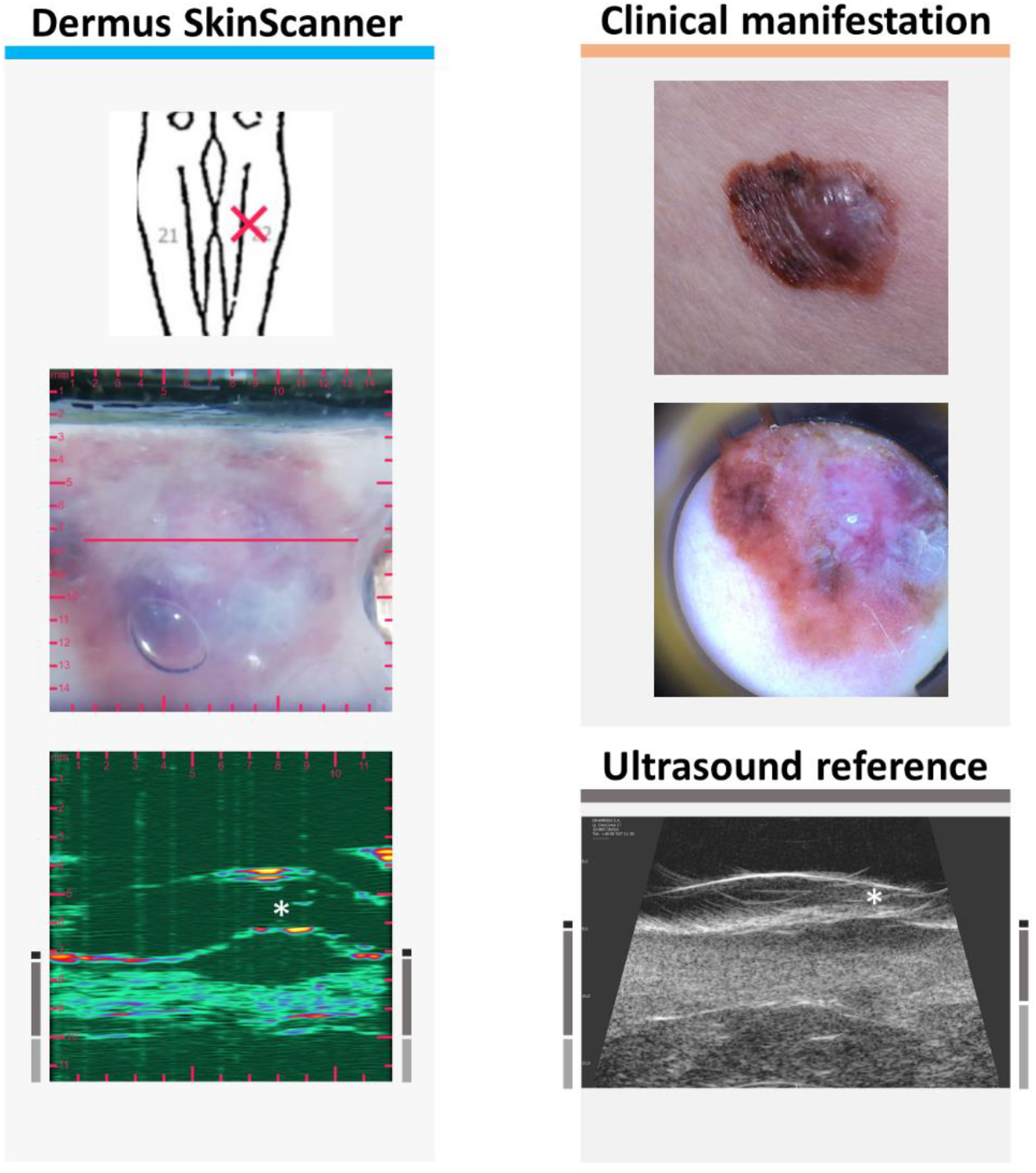
Examination images of a melanoma malignum (see Figure 2 for figure layout information). The ultrasound images show the characteristic hypoechoic spindle-like depth morphology of this melanoma lesion affecting the epidermal and dermal regions.

**Figure 5.**
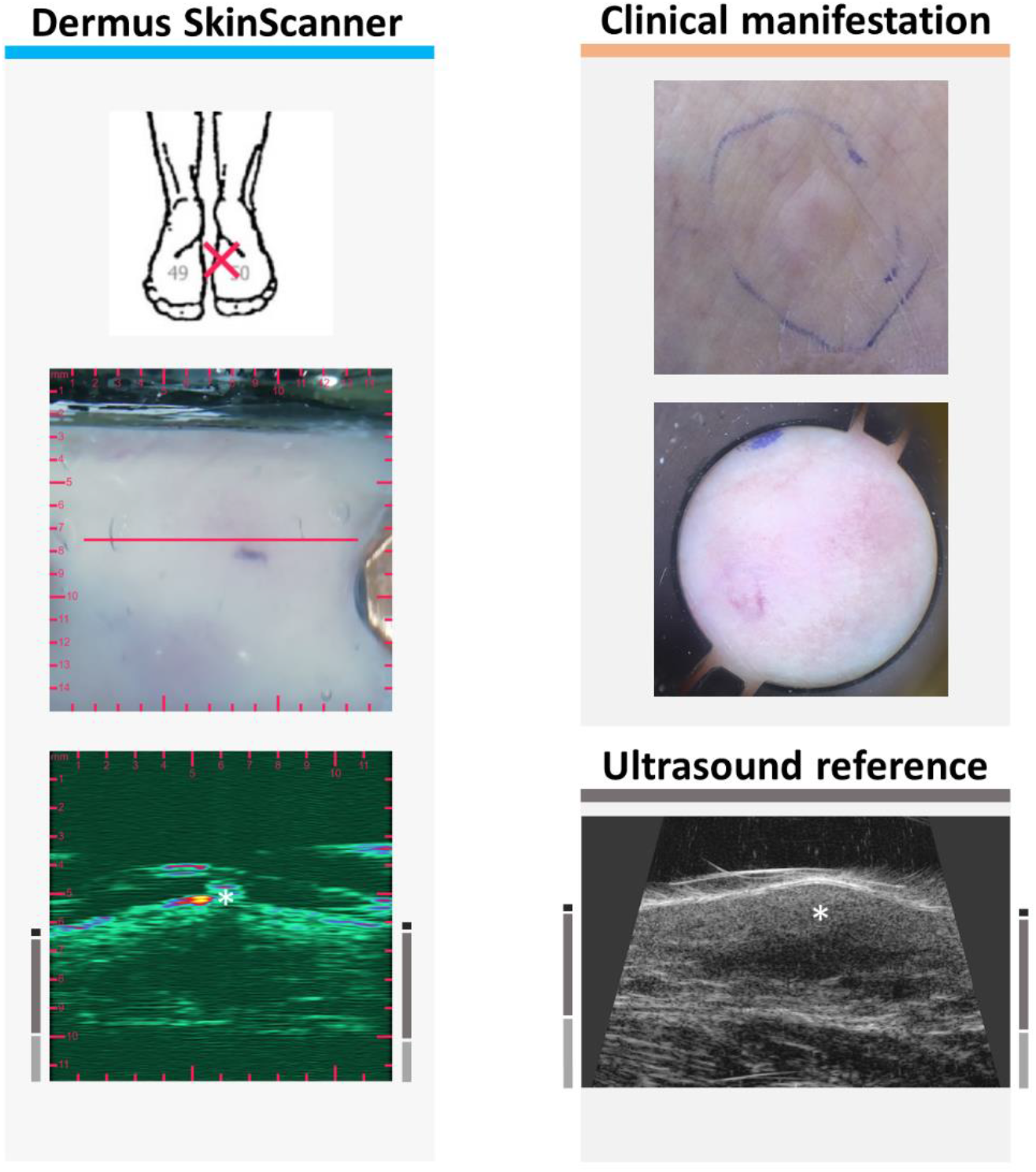
Examination images of a cutaneous metastasis of melanoma malignum (see Figure 2 for figure layout information). No characteristic features can be observed on the optical images of the surface. In contrast, the ultrasound images show a clearly detectable hypoechoic, heterogeneous lesion in the deeper parts of the dermis, with irregular borders and asymmetric shape.

**Figure 6.**
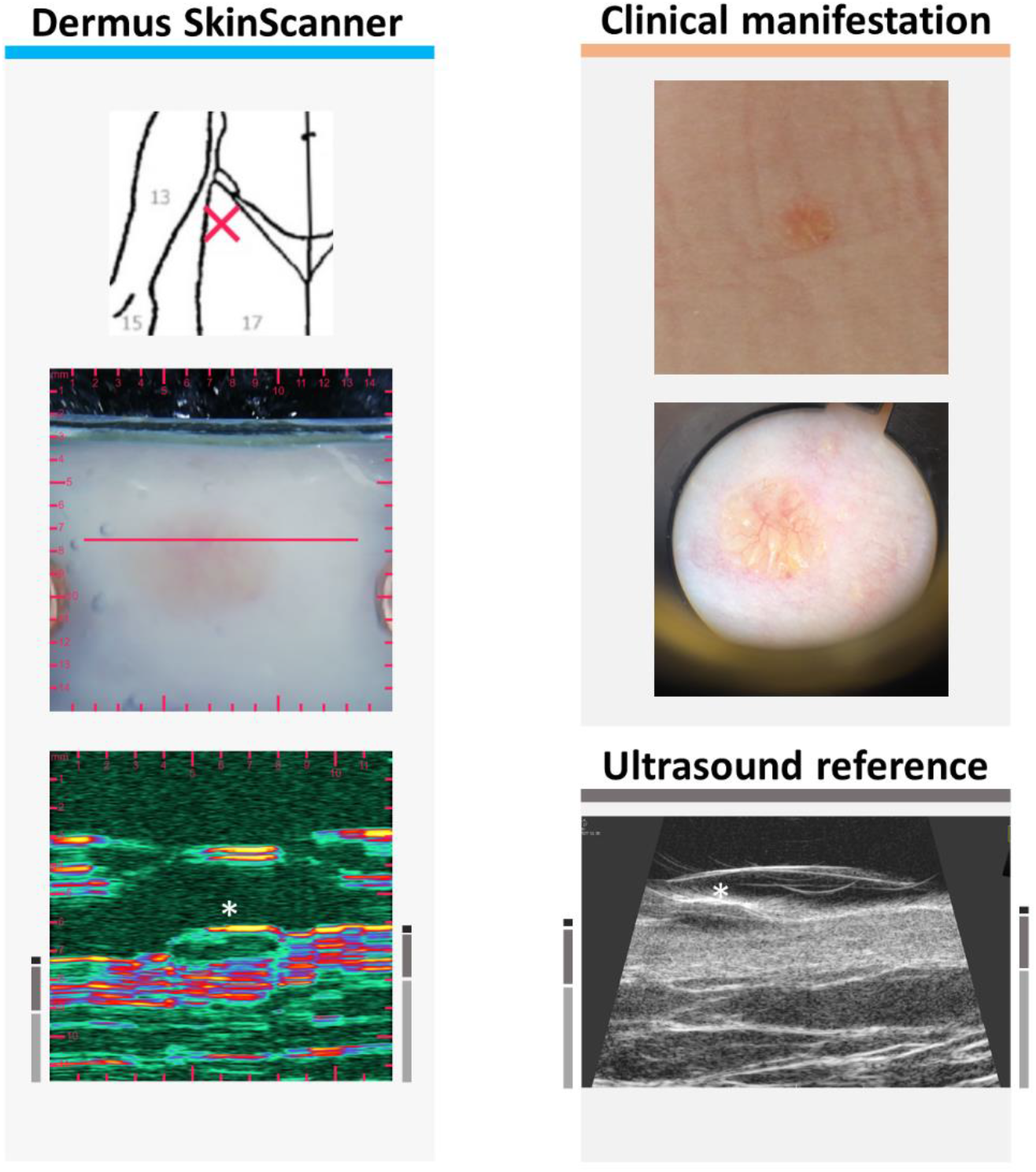
Examination images of a slightly elevated basal cell carcinoma (see Figure 2 for figure layout information). The ultrasound images show a relatively hypoechoic (with echoes from inside), compact but asymmetric lesion. The dermal region of this example is more hyperechoic than those shown in Figures 4 and 5; this represents the variability of skin echogenicity between patients and body locations.

**Figure 7.**
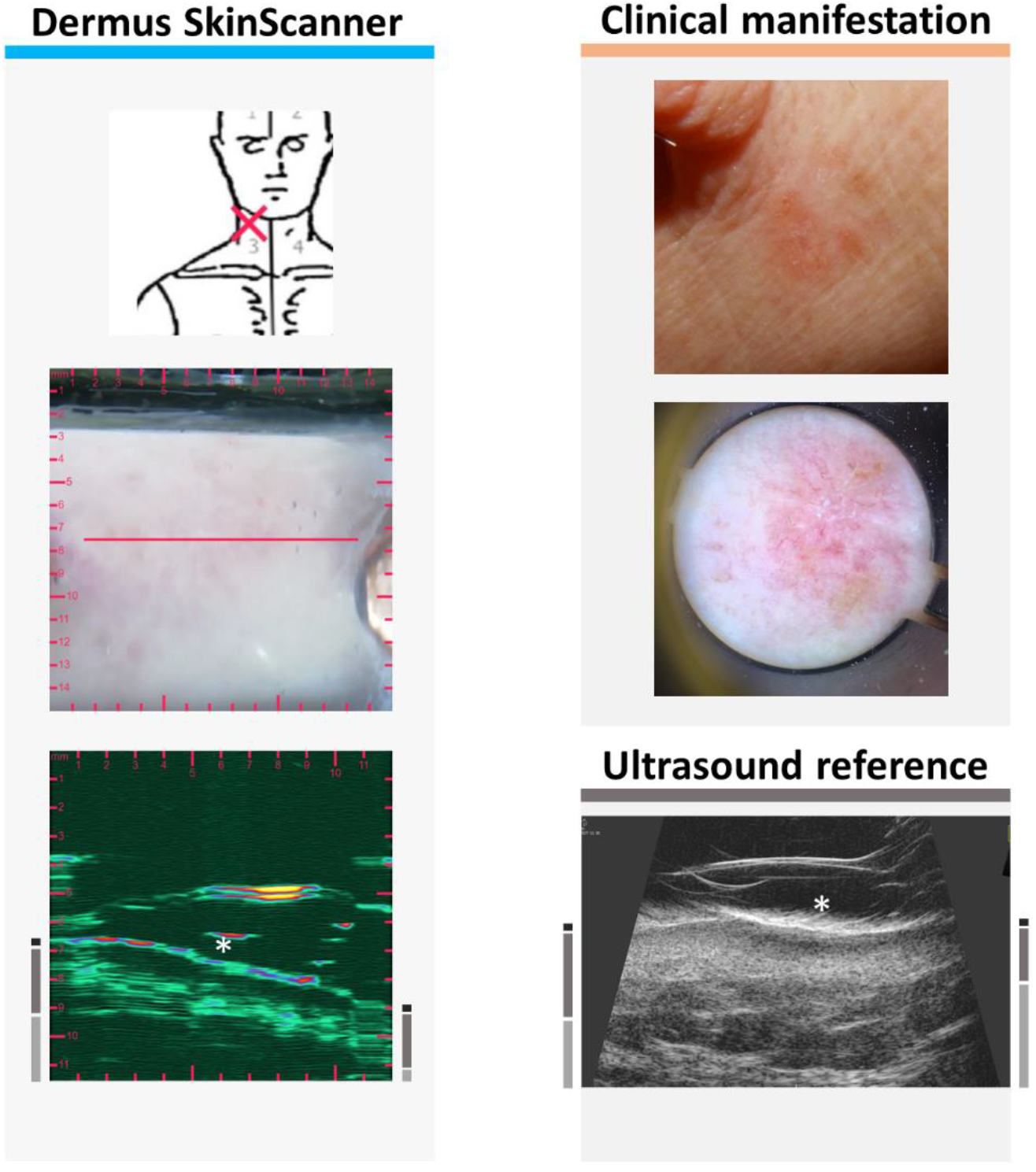
Examination images of a clinically flat, erythematosus basal cell carcinoma that was treated with cryotherapy a year before the examination (see Figure 2 for figure layout information). The ultrasound images show a relatively hypoechoic lesion with hypoechoic boundary regions.

**Figure 8.**
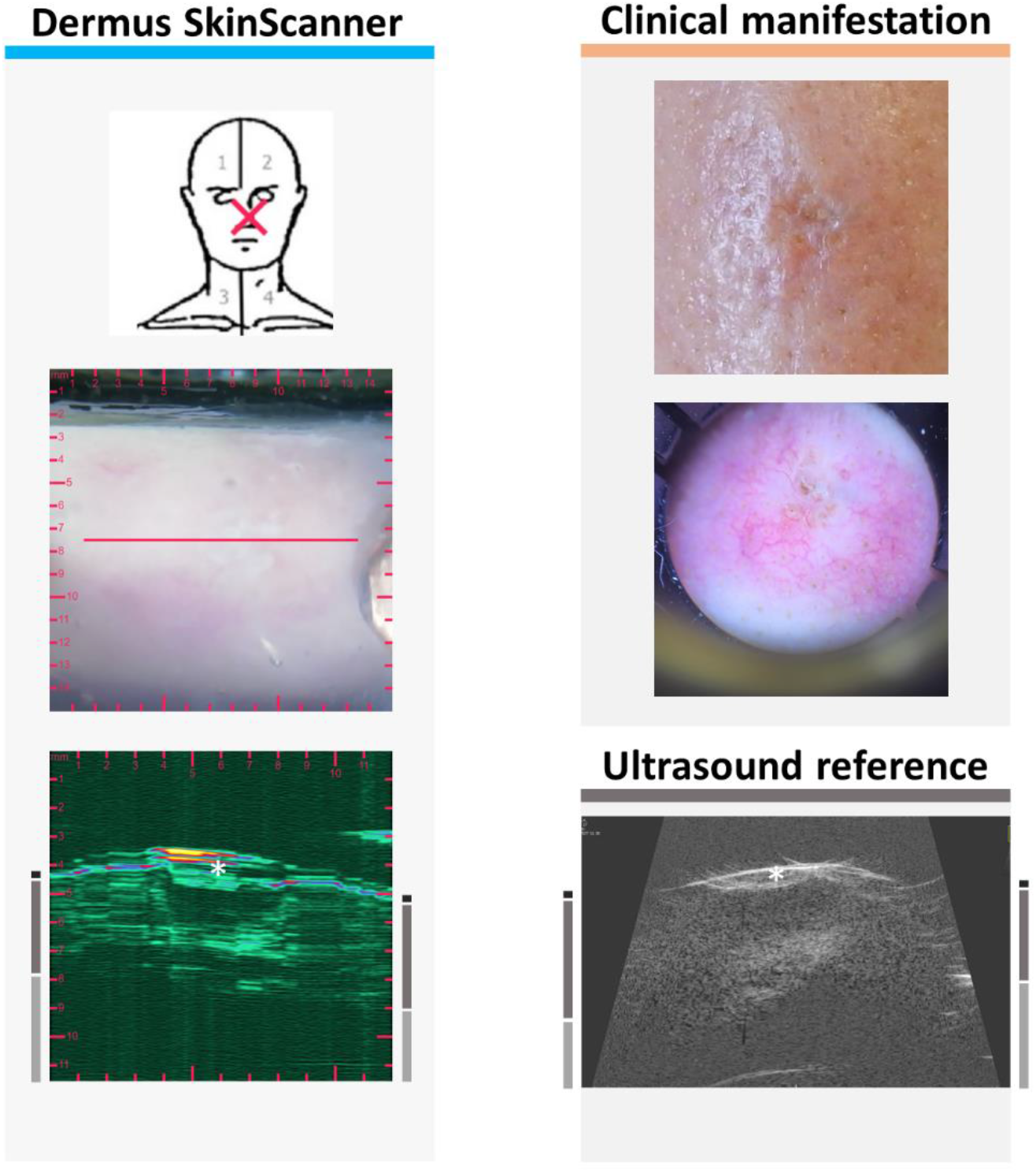
Examination images of a slightly sunken, scaly basal cell carcinoma that was treated with electrocautery therapy 3 months before the examination, being present in a location at which it is relatively hard to perform ultrasound imaging. (See Figure 2 for figure layout information.) The ultrasound images show a heterogeneously hypoechoic lesion, with border irregularities.

**Figure 9.**
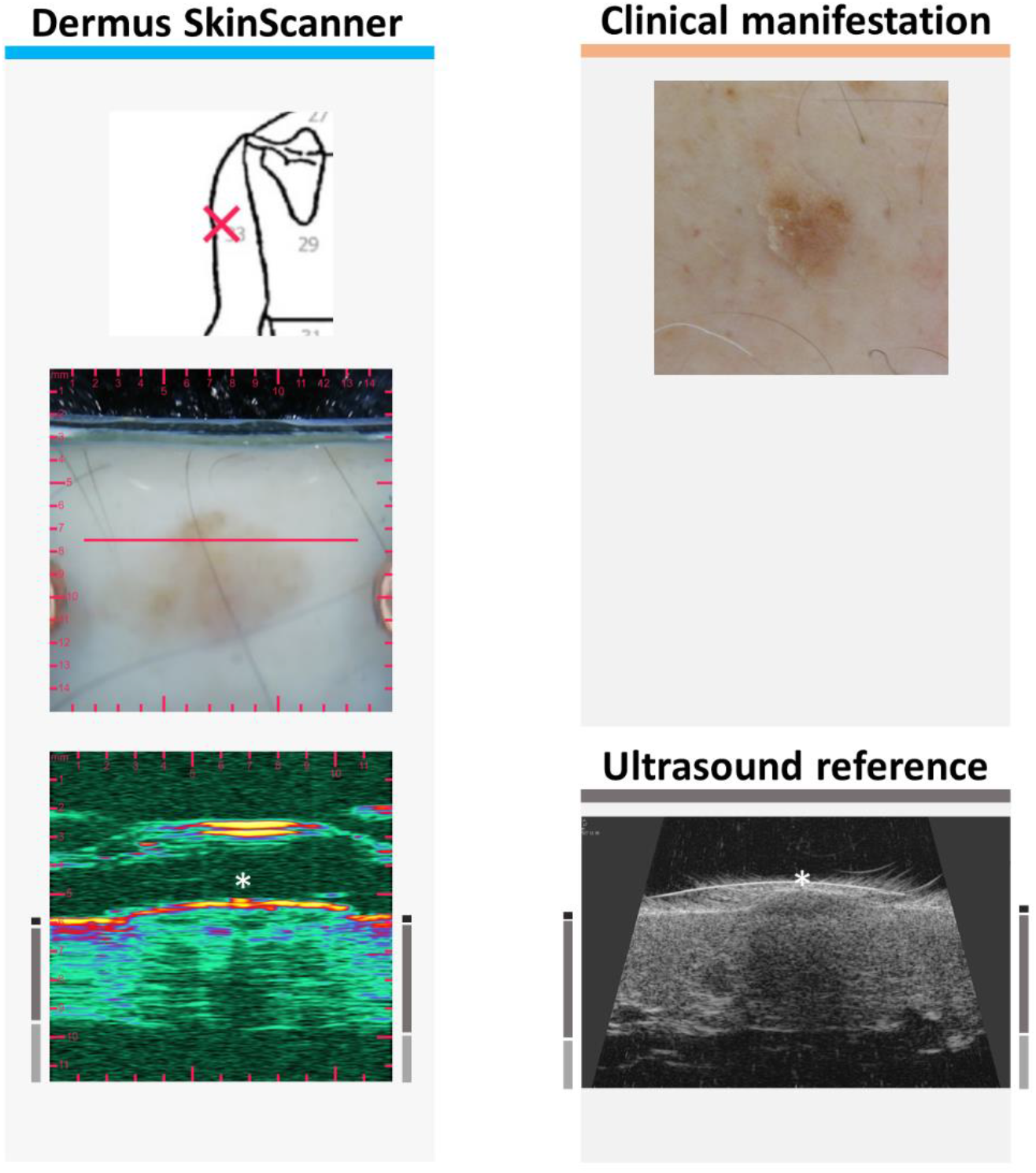
Examination images of a slightly elevated keratotic papule (see Figure 2 for figure layout information). The ultrasound images present the acoustic shadowing effect caused by the relatively strong reflectiveness of the keratotic surface of the lesion.

**Figure 10.**
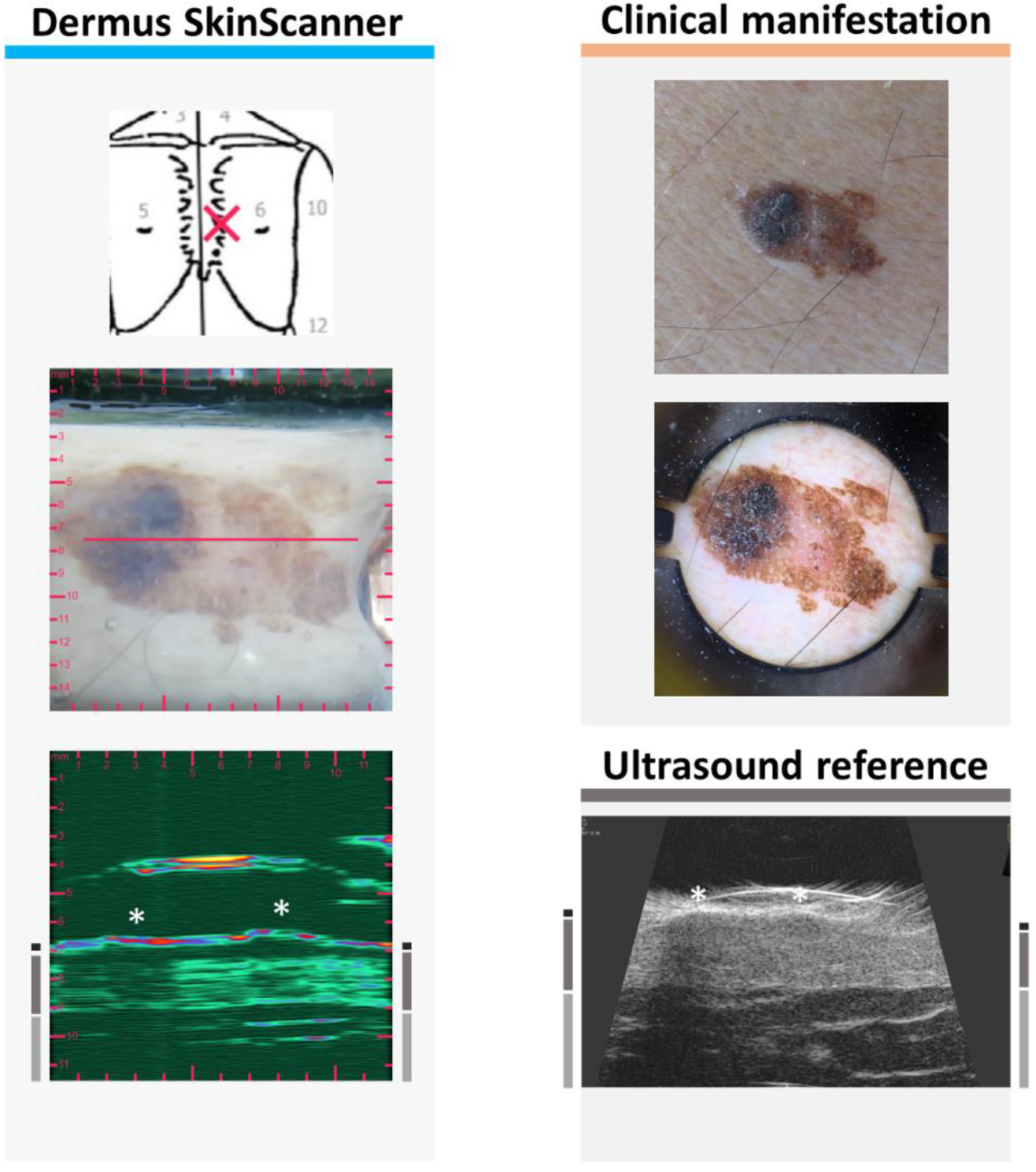
Examination images of a slightly elevated keratosis seborrhoica with asymmetric shape and color (see Figure 2 for figure layout information). The intensity of the acoustic shadows shown on the ultrasound images follow the different scales of reflectivity of the surface of different areas of the asymmetric lesion.

**Figure 11.**
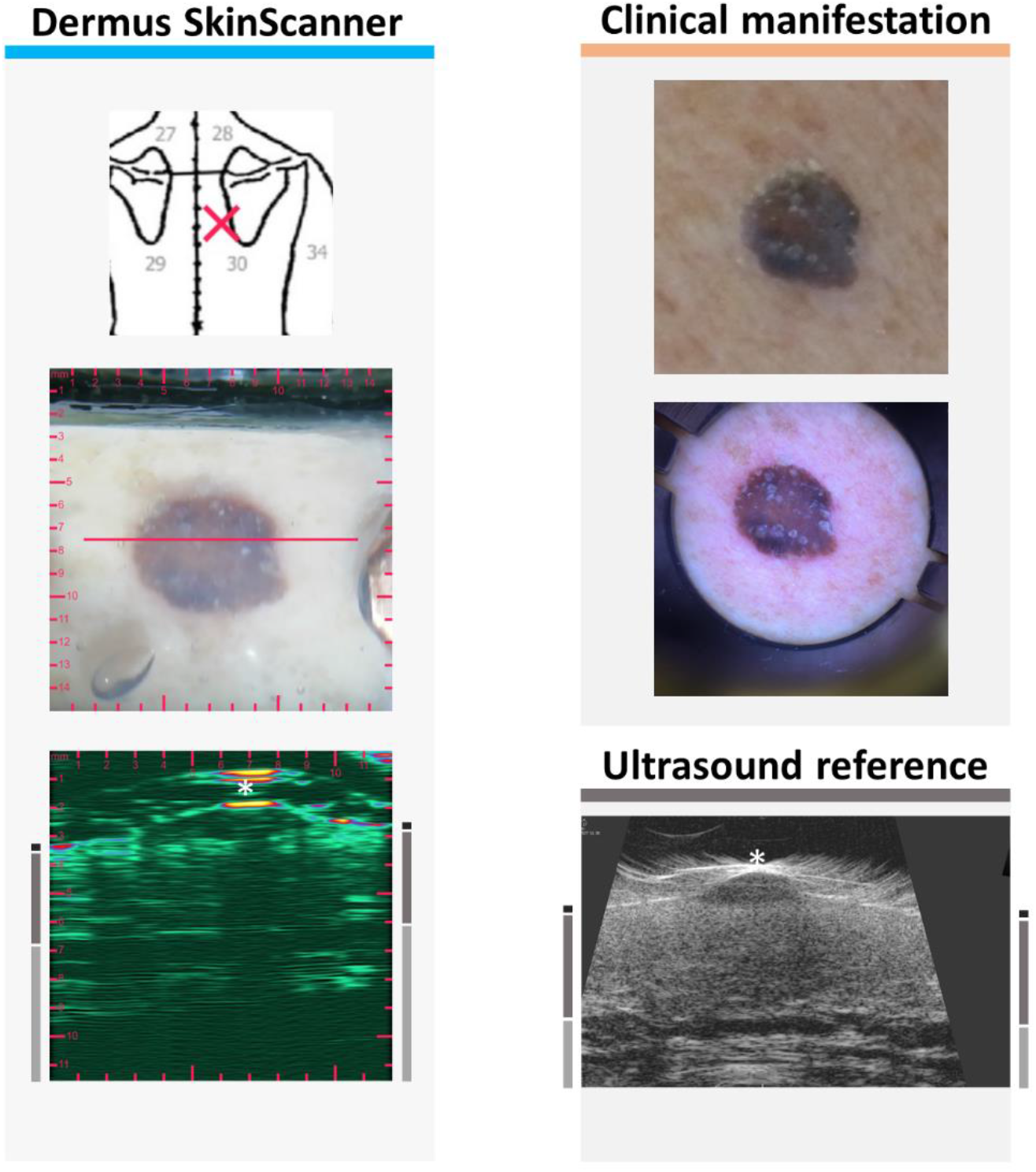
Examination images of a keratosis seborrhoica (see Figure 2 for figure layout information). This lesion showed a suspicious spindle-like subsurface morphology (similar to those of melanoma, see Figure 4) on the ultrasound images, while other features, such as the presence of acoustic shadow, the relatively high echogenicity of the lesion area and the dermoscopic features (sharply demarcated borders, comedo-like openings, cerebriform structures) suggesting the correct diagnosis (keratosis seborrhoica). This example underlines the importance of the holistic, complete evaluation of optical and ultrasound information.

**Figure 12.**
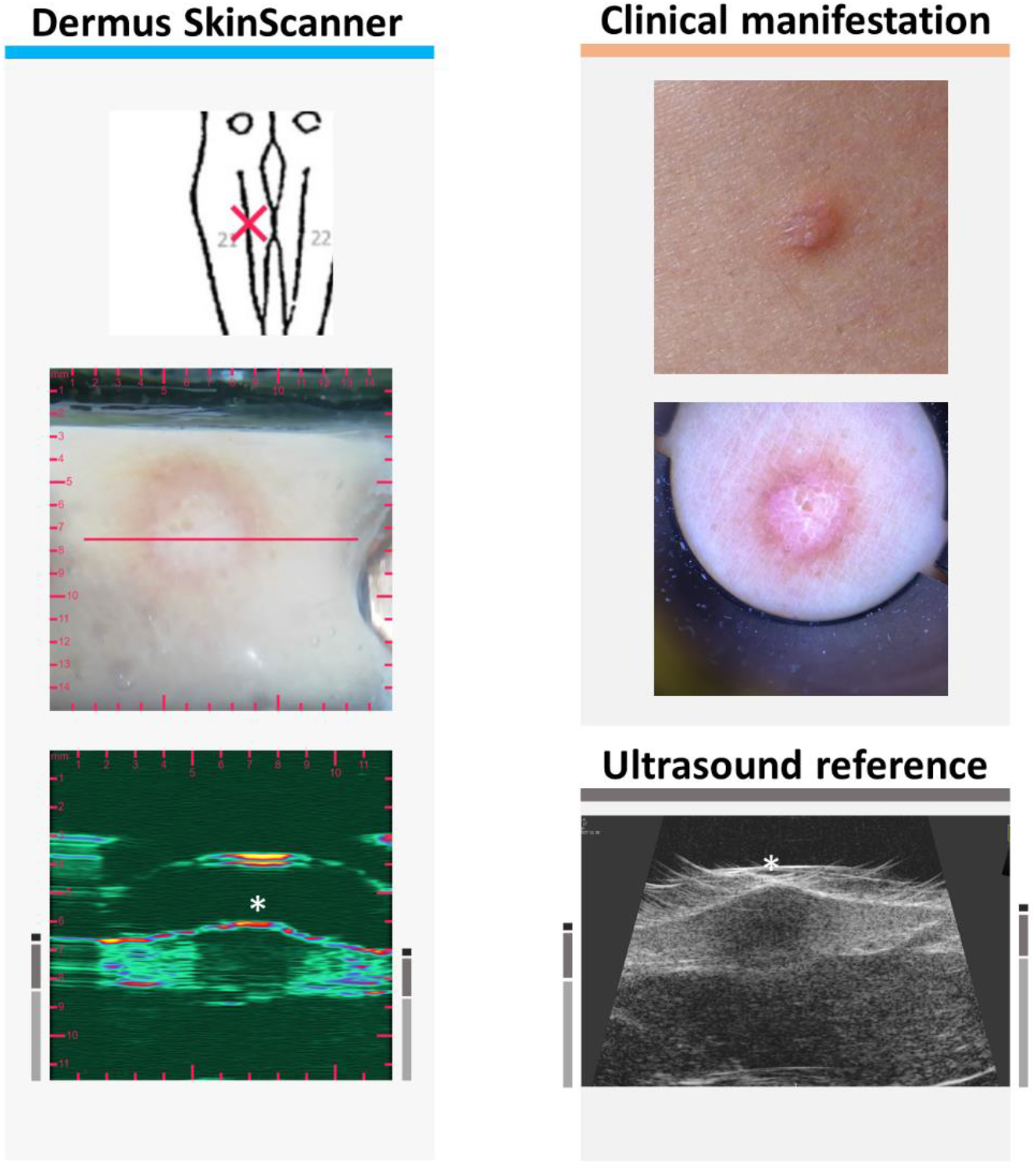
Examination images of a dermatofibroma (see Figure 2 for figure layout information). The ultrasound images show a hypoechoic, heterogeneous lesion in the dermis.

**Figure 13.**
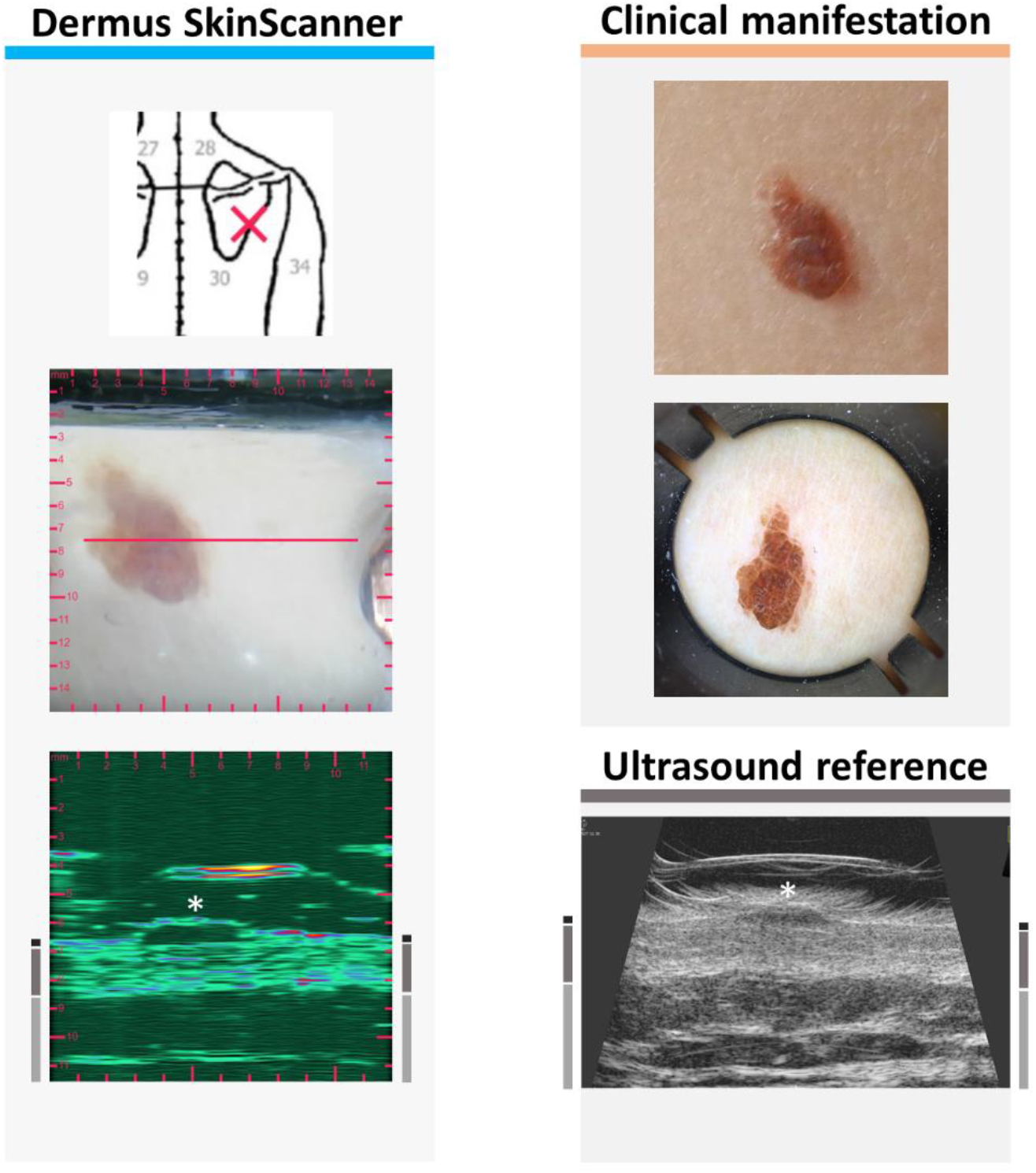
Examination images of an asymmetric brownish naevus (see Figure 2 for figure layout information). The ultrasound images of the lesion show a hypoechoic, well delimited structure with a half-ellipse-like morphology.

**Figure 14.**
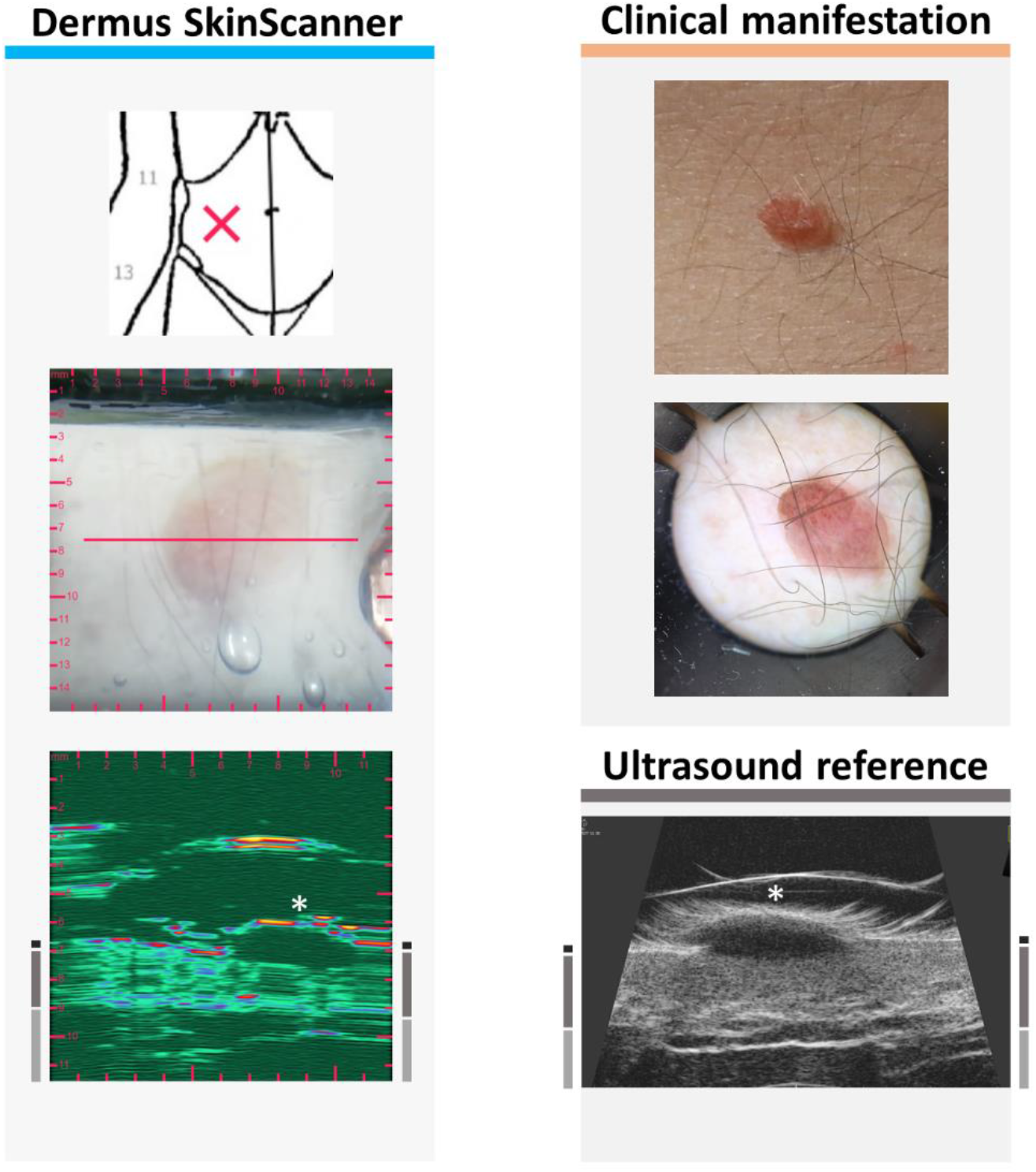
Examination images of a naevus (see Figure 2 for figure layout information). The ultrasound images show a hypoechoic, well delimited half-ellipse-like morphology.

**Figure 15.**
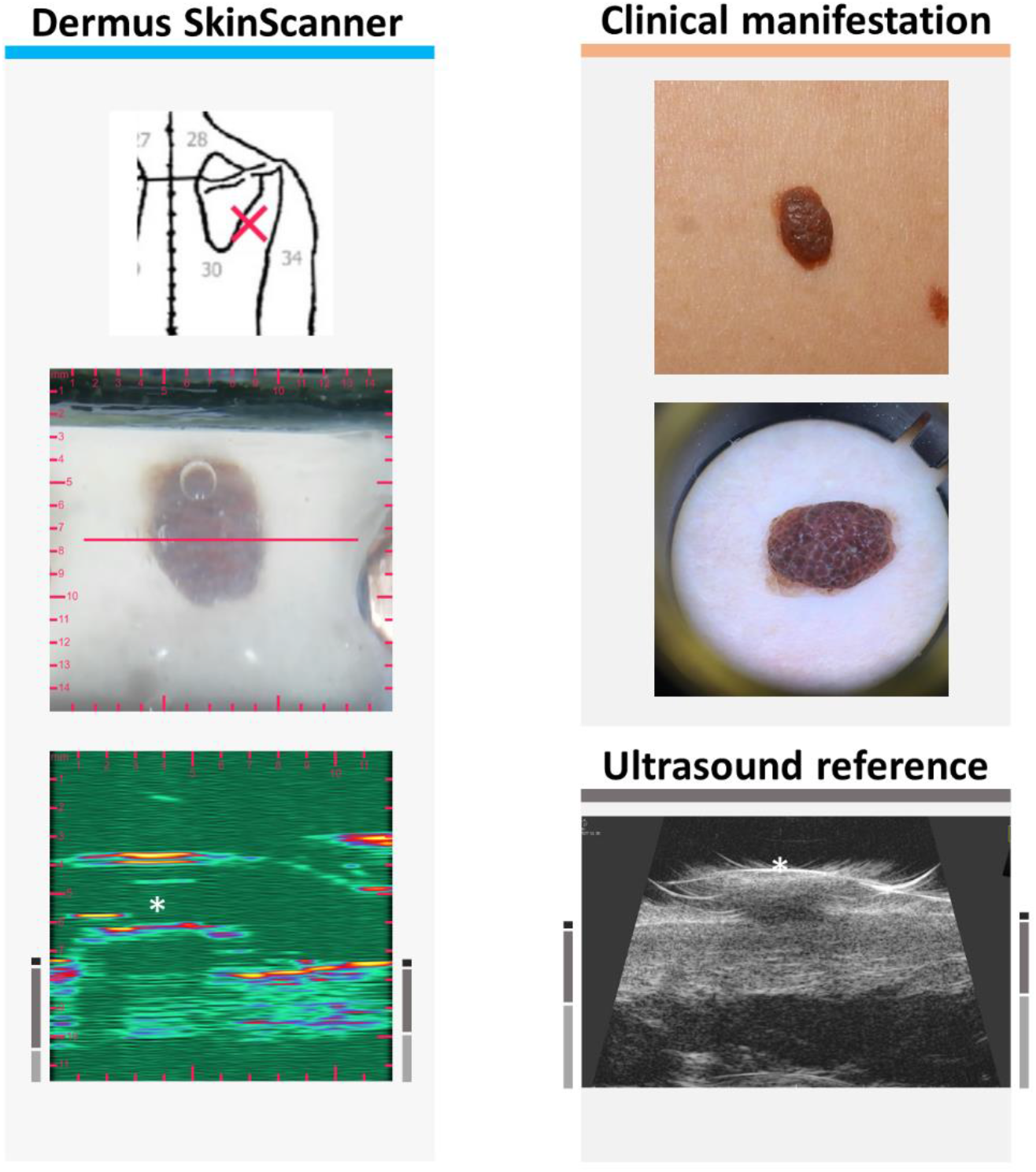
Examination images of a compound naevus (see Figure 2 for figure layout information). The ultrasound images show the relatively hypoechoic, well delimited lesion extending also to the non-elevated area of the dermis. Note that the acoustic shadow on the left of the Dermus SkinScanner ultrasound image is caused by a small air bubble inside the gel in intersection with the red line, right above the left side of the lesion.

**Figure 16.**
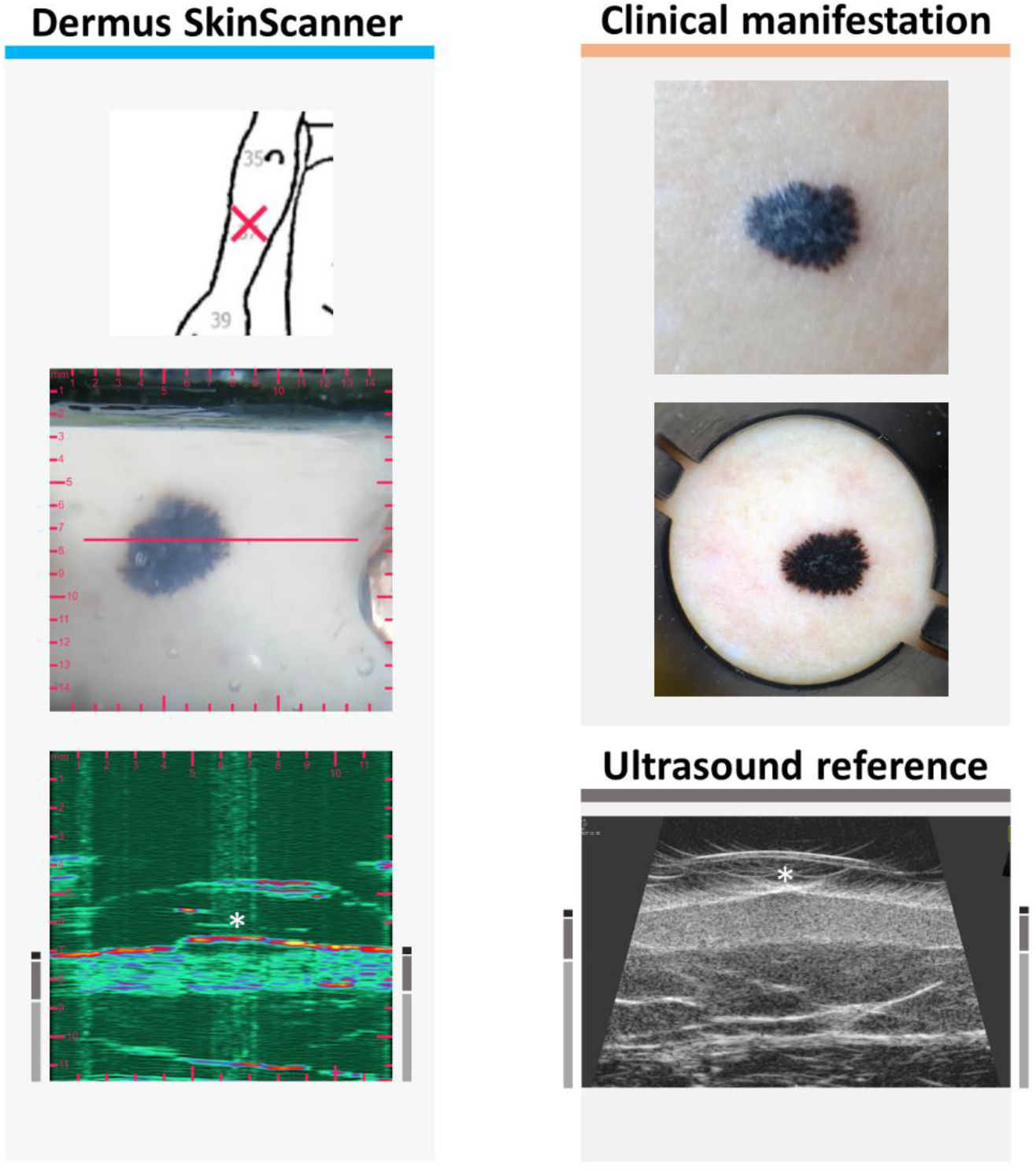
Examination images of a Reed naevus (see Figure 2 for figure layout information). The ultrasound images show the lesion as a hypoechoic structure just below the epidermis.

### 3.1. Skin structure detectability

The results for the detectability of different structures (lesion, epidermis, dermis, subcutis) of the skin, as evaluated by both the investigator and by two independent experts (Section 2.2.3), are summarized in Table 1.

**Table 1.** Detectability of the lesion and primary layers associated with the skin for the investigated and reference devices. (N=53)

| Is the ...<br>distinguish<br>able? | Dermus SkinScanner |  |  | Reference device<br>(Dramiński DermaMed) |  |  |
| --- | --- | --- | --- | --- | --- | --- |
|  | Investi-<br>gator,<br>on site<br>(dermato-<br>logist) | Inde-<br>pendent<br>evaluator<br>(dermato-<br>logist) | Inde-<br>pendent<br>evaluator<br>(radiolo-<br>gist) | Investi-<br>gator,<br>on site<br>(dermato-<br>logist) | Inde-<br>pendent<br>evaluator<br>(dermato-<br>logist) | Inde-<br>pendent<br>evaluator<br>(radiolo-<br>gist) |
| Lesion | 95 % | 91 % | 91 % | 94 % | 87 % | 94 % |
| Epidermis | 96 % | 98 % | 92 % | 96 % | 100 % | 97 % |
| Dermis | 96 % | 91 % | 96 % | 94 % | 96 % | 96 % |
| Subcutis | 94 % | 85 % | 91 % | 94 % | 92 % | 96 % |

On average, the detection performance of the SkinScanner device was 93.0 %, while that of the reference device was 94.7 %, showing the SkinScanner to be non-inferior within a margin of 5% with a confidence interval of 95 %.^28^ The detectability of the lesions seemed slightly better for the *Dermus SkinScanner* images while that of the skin layers seemed superior for the reference device. This was in line with the overall experience description given by the evaluators, where they concluded that the *Dermus SkinScanner* images generally provided better contrast of the lesions with clearer borders, while the reference device was generally more applicable for imaging relatively larger lesions (with >1.5 cm diameter).

The investigator reported being able to distinguish both the lesions and the skin layers on the vast majority (92 %) of the ultrasound images of both devices. In only 4 % of the cases, he reported being unable to distinguish any of these structures on both images (in the cases of a large lesion below the nose, and of a thin lesion directly next to a toenail); in another 4 % of the cases, he was able to distinguish some structures more surely on the *Dermus SkinScanner* images.

Comparison of the evaluations showed a relatively high correspondence (with differences within 10 %) for the two independent and the on-site evaluations. The results suggest that the interpretation of the images of the *Dermus SkinScanner* is slightly aided by relatively minimal training and experience for a non-radiologist user with knowledge of the skin (e.g. dermatologist).

### 3.2. Dermus SkinScanner ultrasound image interpretation

The *Dermus SkinScanner* displays 2-D ultrasound images with a color code for a greater contrast between the structural elements on the images. A dark color represents the lowest intensity which is followed by green and blue up to red and yellow which stands for the highest intensity value.

Regarding the structures of the skin, ultrasound reflections from the epidermis usually appear as a high-intensity, thin structure on the images. The dermis appears as a thicker structure with inhomogeneities in ultrasound reflection intensity. The subcutis usually appears as a low-intensity region with scattered structures in it appearing with a higher intensity. This is consistent with the literature on other dermatological ultrasound images.^2^

Typical elements of the ultrasound images of the *Dermus SkinScanner* are presented in Figure 3 using representative images of a cutaneous inflammation and of a cutaneous neoplasm. The spatial location of an ultrasound image slice is indicated by the red line on the corresponding optical image of the surface.

It should be noted that several artefacts may appear on the images besides the structures of interest. These artefacts include the ultrasound reflections from the outer structures and from the membrane of the imaging window, possible air bubbles of the gel and consequent acoustic shadows on the image of the deeper structures below the bubbles. Regarding the reflections related to the imaging window, the artefacts are planned to be removed or at least substantially reduced for an easier interpretation of the images in the next generation of the device. Regarding air bubbles, the related artefacts can be reduced or eliminated with special attention when applying the gel on the surface of the imaging window.

### 3.3. Melanoma

Figure 4 presents a typical case of melanoma malignum. Clinically, on the surface, the lesion appears with asymmetric shape and diverse colors, with a central elevated part. The dermatoscopy shows a disorganised pattern, asymmetric pigmentation, with multiple colors, blue-whitish veil and atypical vessels. Regarding the ultrasound images, the characteristic hypoechoic spindle-like depth morphology of malignant melanoma is clearly observable in Figure 4. The thickness of the lesion was measured to be 1.97 mm on the ultrasound image of the *Dermus SkinScanner*. This is slightly higher than the 1.68 mm thickness measured by histopathology, as expected due to tissue shrinkage after excision and during histological preparation.^29^ The thickness measured on the reference ultrasound image, however, is smaller than both the histopathological and *Dermus SkinScanner* measurements: 1.17 mm. This could be due to the difficulty of imaging the right cross-section of the lesion on this image without an optical aid.

It is significantly more difficult to identify a cutaneous metastasis of melanoma since it has no characteristic clinical and dermatoscopic features. However, Figure 5 shows that its presence is clearly noticeable and localizable on the ultrasound images. It appears as a hypoechoic, heterogeneous lesion in the deeper parts of the dermis, with irregular borders and asymmetric shape. These observations are in correspondence with those found in the literature.^30^

### 3.4. Basal cell carcinoma

Basal cell carcinomas (BCC) may not show clear borders as seen from the surface of the skin, therefore skin ultrasound examination can help in clarifying these borders.^21,31^ This makes the ultrasound examination of BCCs particularly relevant.

In Figure 6, a BCC is present optically as a shiny, slightly elevated yellow-whitish papule, dermatoscopy shows arborising vessels and shiny white blotches. Regarding the ultrasound images, a relatively compact but asymmetric morphology is present (being distinguishable from the spindle-like morphology of Figure 4). The lesion is generally more hypoechoic than the surrounding dermis but echoes of ultrasound signals can be observed within the lesion (it is less hypoechoic than a typical melanoma). It is observable on both ultrasound images that the skin area presented in this case contains a dermis that is relatively highly hyperechoic.

Figure 7 presents a clinically flat, shiny erythematosus lesion, with dermatoscopy showing arborising vessels, shiny white blotches and signs of ulceration. The ultrasound images show a relatively hypoechoic lesion with a more hypoechoic central region and less hypoechoic boundary regions, and with a relatively irregular morphology.

There are body locations in which ultrasound imaging is relatively more difficult to perform – such as inclinations and curves of the nose, ears, eye area, mouth area. It is presented below in Figure 8 that it is possible to record relatively good quality images even in those areas.

The BCC presented in Figure 8 is clinically a shiny, slightly sunken, scaly lesion. Dermatoscopy shows arborising vessels and ulcerations. A heterogeneously hypoechoic lesion, with a more hypoechoic center and less hypoechoic boundary regions with border irregularities, is present in the dermis on the ultrasound images. The lesion borders are evidently more detectable on the *Dermus SkinScanner* images as compared to the reference image in this case.

### 3.5. Keratosis

Figure 9 depicts a clinically slightly elevated, brownish keratotic papule. In comparison, Figure 10 depicts a clinically asymmetric shape and color, with a slightly elevated, keratotic surface, with the dermatoscopy showing sharply demarcated borders, milia-like cysts, fissures and ridges, and network-like structures.

In terms of ultrasound features, ultrasound images of keratoses often include some acoustic shadows caused by the relatively strong reflectiveness of the keratotic surfaces to the ultrasound waves. Accordingly, strong reflections from the uppermost layer of the lesions and the shadows beneath this layer may be noticed on Figures 9–10. Figure 10 presents an example of a keratosis seborrhoica with parts of its surface showing different scales of reflectivity that is followed by the intensity of the acoustic shadows on the corresponding ultrasound images.

An interesting case is presented in Figure 11. Clinically, shape and color asymmetry are visible. The dermoscopy image shows sharply demarcated borders, comedo-like openings and cerebriform structures. The ultrasound images of the depth slice of this lesion showed a suspiciously spindle-like morphology suggesting that it may be a melanoma with an atypical surface image. However, other features (such as the presence of a moderate keratosis-like shadowing and as the dermoscopy features) were suggesting it being a keratosis seborrhoica which was then confirmed by final (histopathological) diagnosis.

### 3.6. Dermatofibroma

Figure 12 presents the images of a dermatofibroma (histiocytoma). Clinically it shows as a firm pink papule with a pale central part. The dermatoscopic image is characteristic showing a central whitish-pink area surrounded by a faint brownish pigmentated network. On the ultrasound images, it appears as a hypoechoic but heterogeneous lesion in the region of the dermis, with unclear borders.^32^

### 3.7. Naevus

The naevus of Figure 13 was introduced to the clinical examination as an asymmetric brownish lesion. Its dermatoscopic image shows a homogenous structure with no sign of malignancy. The corresponding ultrasound image presents the lesion as a hypoechoic, well demarcated dermal structure with a half-ellipse-like morphology. The information of the ultrasound image further supported the diagnosis of the dermatoscopic examination.

Another lesion is presented in Figure 14 that is clinically a uniform brown, slightly elevated naevus. Dermoscopy image shows a slight asymmetry in its pigmentation. On the ultrasound image, the signs of benign naevi can be observed, such as the hypoechoic, well delimited half-ellipse-like morphology.

Figure 15 presents a compound naevus. Clinically dark brown elevated lesion, dermatoscopy image shows a papillopmatosus dark-brown central area, with globular pattern; the central part is surrounded by a light brown area. On the ultrasound image, the depth extent of the compound naevus is well visible. Morphologically, the borders and shape on the ultrasound image are not as regular as those of the cases of Figures 13–14, but the combined opto-ultrasonic information gives confidence in the diagnosis.

Figure 16 shows the images of a lesion that was primarily suspicious for being a melanoma, but histopathology proved the diagnosis of a Reed naevus. Clinically, it appears as a dark brown to black, slightly asymmetric pigmented papule, while dermatoscopy shows a dark brown to black color, typical starburst pattern, structureless central area, radial lines and pseudopods distributed symmetrically around the lesion. In the ultrasound images, the lesion presents just below the epidermis in the upper dermis as a hypoechoic structure.

### 3.8. Dermatitis

Inflammatory skin diseases such as atopic dermatitis can also be followed using ultrasound imaging. It was shown in the literature that the severity of the inflammation correlates with the thickness of a subepidermal low echogenic band (SLEB) on their skin ultrasound images.^10,33^ A case of atopic dermatitis is presented on Figures 17–18. Clinically dry skin, mild erythema with erosions and signs of lichenification are visible. The SLEB can clearly be observed on the ultrasound images of Figure 17.

**Figure 17.**
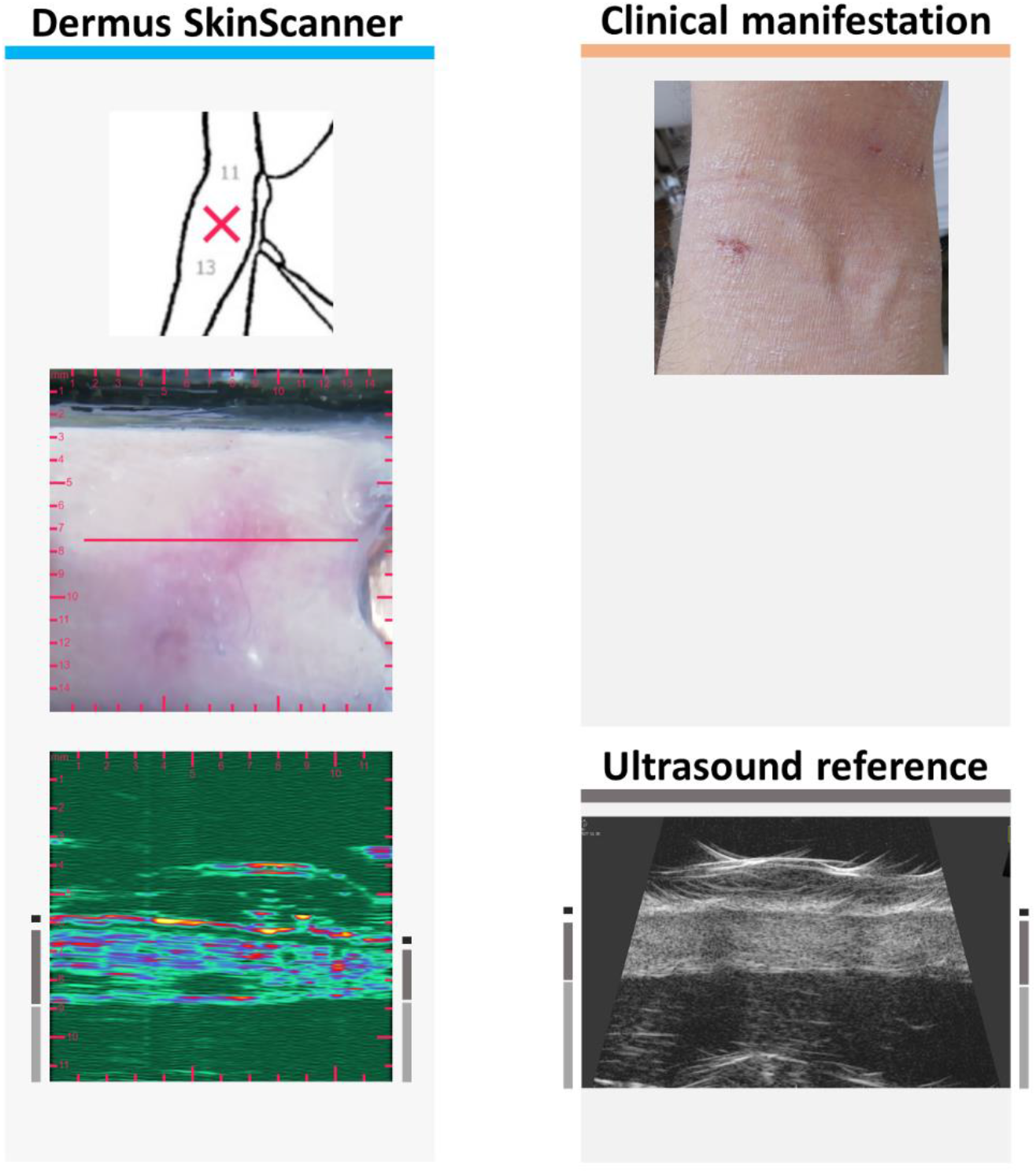
Examination images of inflammatory skin lesion due to atopic dermatitis (see Figure 2 for figure layout information). The SLEB (subepidermal low echogenic band) can be seen on both ultrasound images.

As this patient appeared again in the study, a follow-up examination could be made which is presented on Figure 18 comparing the images of Figure 17 (T1) to the follow up, after two weeks of topical treatment (T2). The differences are clearly detectable when comparing the corresponding images of before (T1) and after (T2) the treatment. On the surface, the erythematosus appearance of the skin has disappeared, and its dryness was significantly reduced. The SLEB present on the ultrasound images of the inflammated skin (T1) has disappeared after treatment (T2). The relative changes in ultrasound reflectivity of the epidermis and dermis, as observed after the treatment, was due to the increased hydration of the skin due to the topical treatment. It is worth noting that there is a detectable difference also in the thickness of the dermis: the dermis was thicker during the inflammation (T1) then after the treatment (T2).

**Figure 18.**
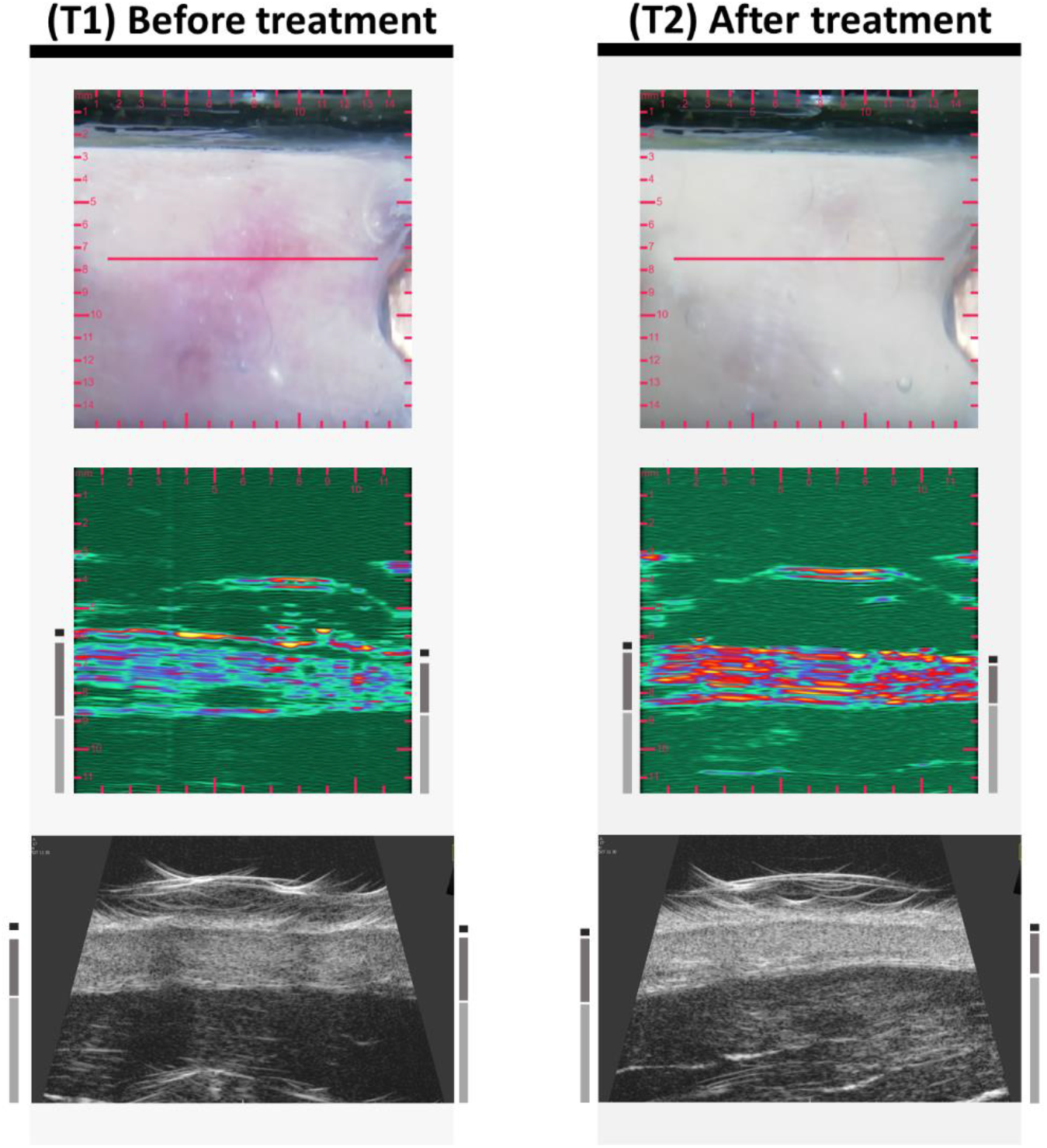
Examination images of the skin area of Figure 17 before (T1) and after (T2) two weeks of topical treatment of atopic dermatitis. A distance of 1 mm is denoted by red tick marks on the SkinScanner optical and ultrasound images and by grey tick marks on the left side of the reference ultrasound images. Shrinking of the SLEB as well as changes in echogenicity of the epidermis (decreased echogenicity) and dermis (increased echogenicity), due to an increased level of hydration, and slight shrinking of dermis thickness can be observed on the ultrasound images of T2 in comparison to those of T1.

### 3.9. Psoriasis

The erythematosus skin area of Figure 19, without significant scaling, is representing an inflammated psoriatic plaque. A clear sign of the inflammation is visible on the ultrasound image as a hypoechoic band beneath the epidermis, presenting the same SLEB ultrasound feature as observed in the cases of dermatitis (Section 3.8). Also note the relatively large thickness of the dermis as another sign associated with the inflammation.^13,34,35^

**Figure 19.**
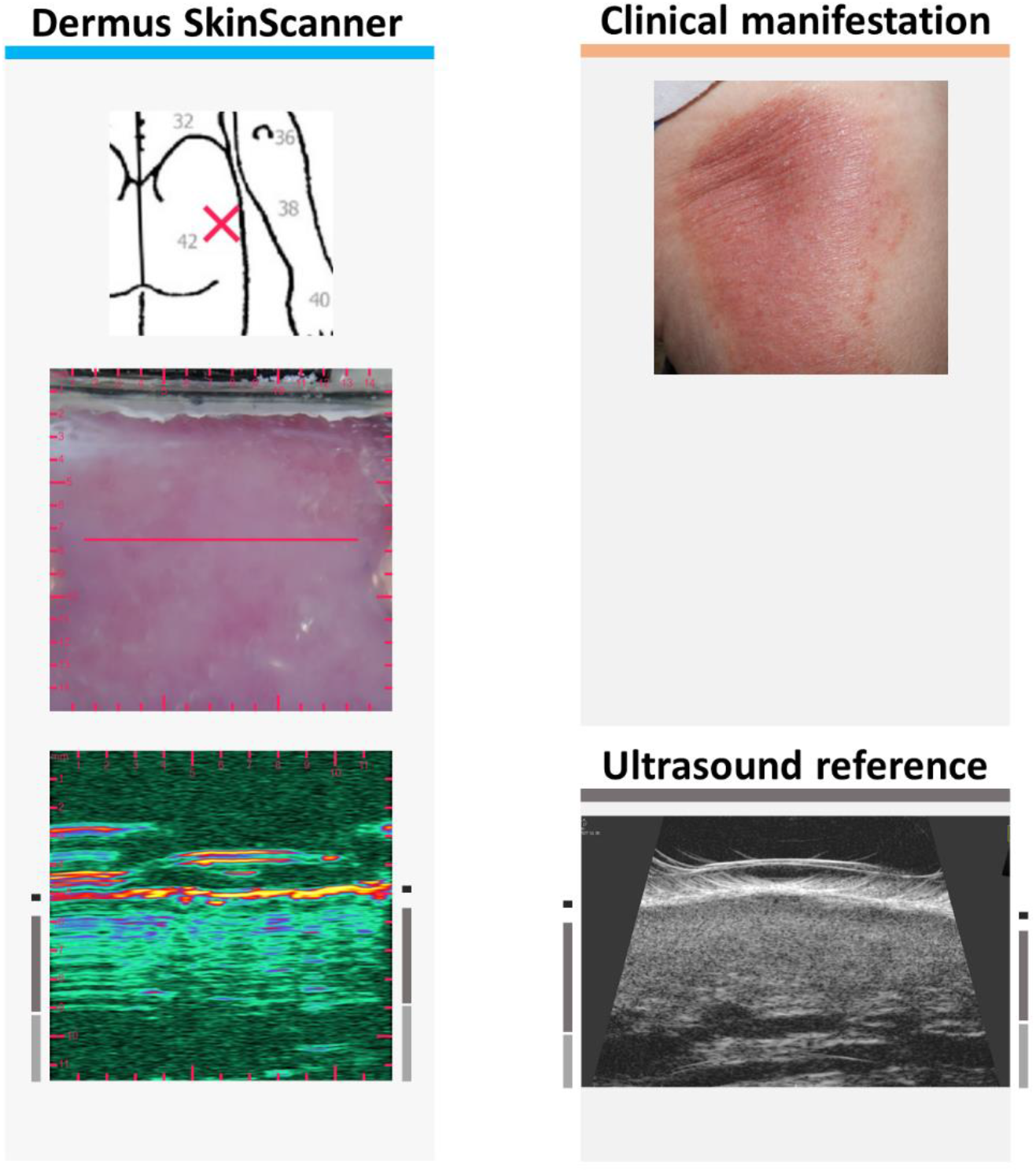
Examination images of inflammatory skin lesion due to psoriasis (see Figure 2 for figure layout information). The ultrasound images show a clearly visible SLEB as well as a relatively high thickness of the dermis (as signs of inflammation).

Figure 20 presents another case of an inflammatory skin lesion due to psoriasis, on the distal end of the left ventral forearm. The lesion appears as a 4–5 cm diameter erythematosus, slightly scaling plaque on its clinical image. On the dermoscopic image, it appears as a vivid erythematosus surface, with tiny erosions, and whitish scales. Both the clinical and dermoscopic images bear the signs of inflammation. The inflammatory skin lesion seen on the optical images is also observable on the ultrasound image with a clearly distinguishable SLEB, that is in correspondence with the observations found in the literature.^13,34^ It is interesting to note that a vessel-like hypoechoic structure in the dermis is also catched in parallel orientation on the ultrasound image presented here.

**Figure 20.**
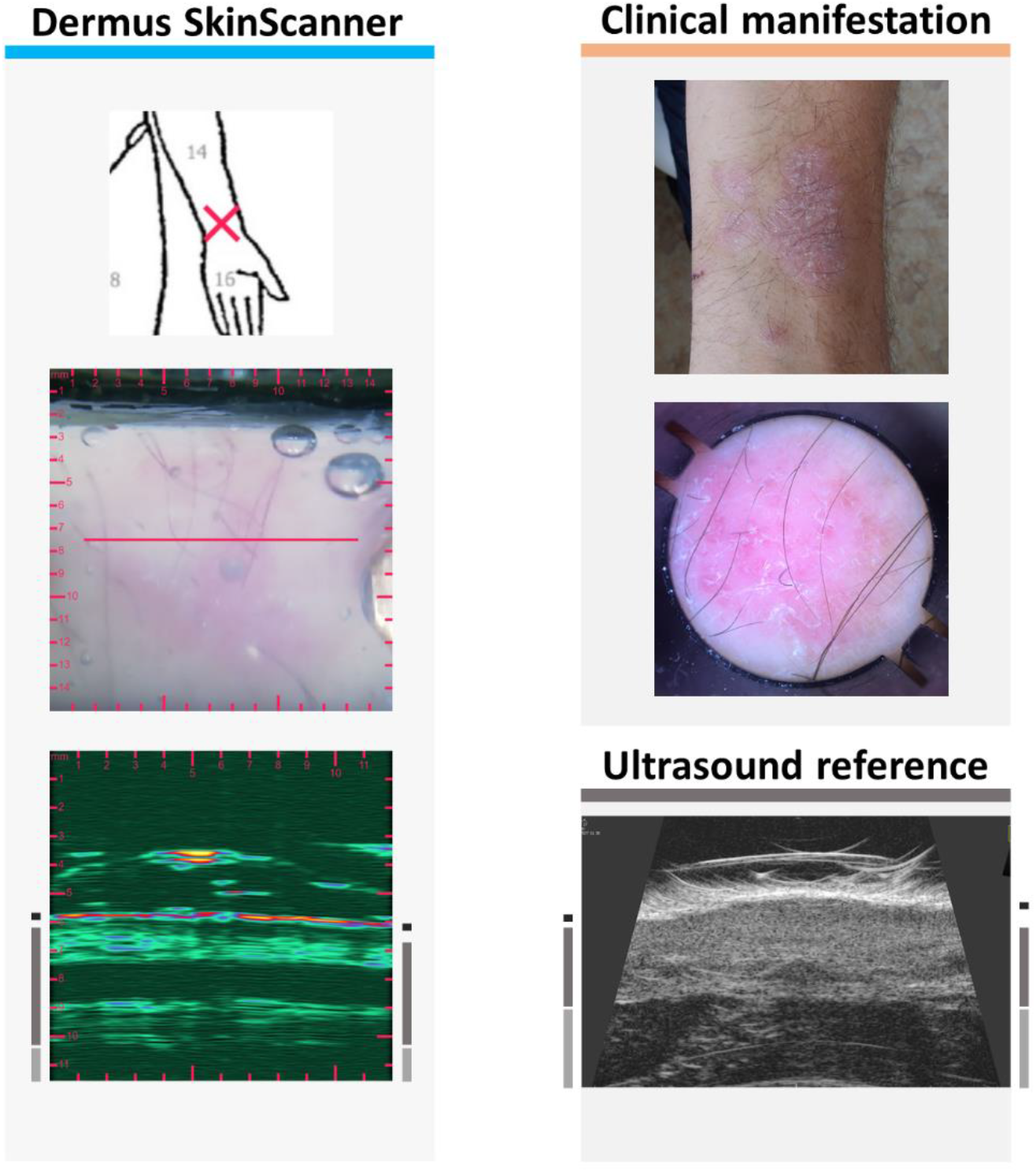
Examination images of inflammatory skin lesion due to psoriasis (see Figure 2 for figure layout information). The ultrasound images show a clearly visible SLEB and a relatively thick dermis. Note the vessel-like hypoechoic structure in the dermis catched in parallel orientation to the image plane.

Figure 21 presents the case of a guttate psoriasis. Small, erythematosus, slightly shiny papules of 0.5–1 cm diameter are being present clinically, with signs of scaling. Dermoscopy shows whitish scales, a small number of red dots is present. SLEB can be clearly distinguished as a marker of inflammation, on the ultrasound images. It is not that spacious as on the images of Figures 19–20 due to the guttate nature of the inflammation.

**Figure 21.**
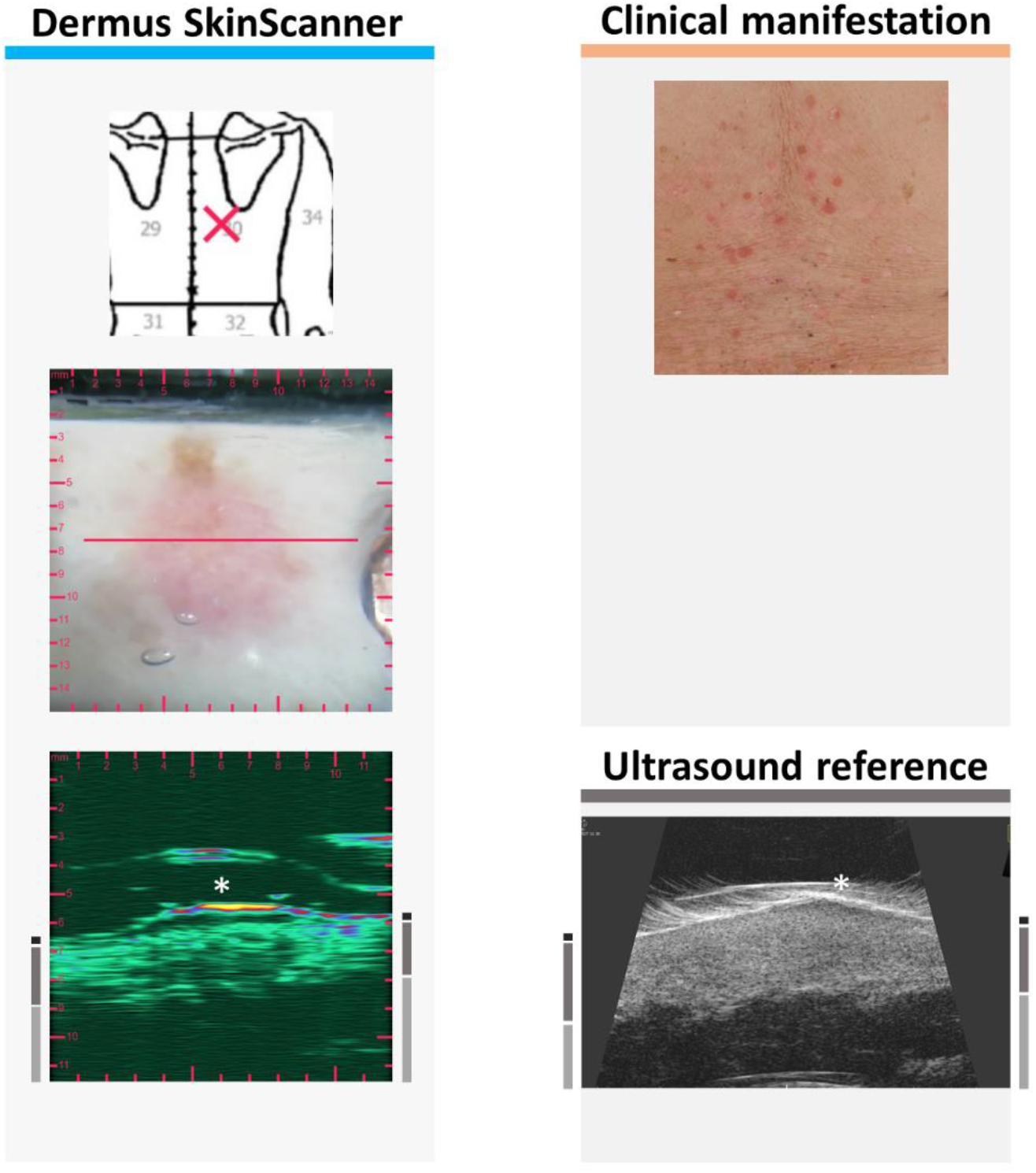
Examination images of inflammatory skin lesion due to guttate psoriasis (see Figure 2 for figure layout information). A SLEB with a relatively small lateral extent (compared to those of Figures 17, 19, 20) is shown on the ultrasound images.

## 4. Discussion

As described in the Results section (Section 3.1), the *Dermus SkinScanner* gave comparable skin structure detectability to the reference device, with experience in using the devices seemingly giving some advantage in interpreting the images. When comparing representative images of different skin diseases, images from both ultrasound devices showed similar features. These features are also consistent with those documented in literature, as discussed below.

Both devices showed malignant melanoma with a spindle-like, fusiform shape. This is one of the typical shapes that is also reported in the literature,^36-39^ one that may be used to distinguish it from other skin lesions. The irregular shape of the metastatic melanoma recorded in the current study has also been noted in the literature.^37^ Since such metastases cannot be seen on the surface they present a unique opportunity for the ultrasound to provide diagnostic assessment.^40^

In the case of the basal cell carcinoma, several of the recordings showed hyperechoic spots within the lesion (Figures 6 and 8); this feature is noted in the literature as being one of the characteristic signs of BCC,^22,36,37,39-41^ which could therefore potentially be used to differentiate the lesion from other lesion types. In addition, since the outline of BCCs is difficult to assess optically, ultrasound may serve as an aid in treatment planning.^42^

Although less reported in the literature, the features observed on the keratoses, dermatofiborma and naevi also corresponded with that observed in the literature. The strong reflectivity of the keratotic surface leads to shadows underneath.^41,43^ Literature expects the dermatofibroma lesion to be ill-defined and heterogeneous, which corresponds to the ultrasound images shown on Figure 12. The naevi in Figures 13 and 14 are well-delimited and hyperechoic as reported in.^40^

Finally, skin inflammations such as atopic dermatitis and psoriasis typically present a subepidermal low echogenic band (SLEB)^22^ whose thickness can be related to the diseases severity.^22,44,45^ Interestingly, the reaction to therapy can also be observed both in psoriasis^40,41^ and dermatitis,^12^ as has also been observed in the current study, where a shrinkage of the SLEB occurred as a response to treatment of atopic dermatitis, as well as changes in the echogenicity of the epidermis and dermis, and dermis thickness.

So far, it has been demonstrated that the features observed on images recorded by the *Dermus SkinScanner* as well as by the reference device correlate well with those observed in the literature. However, the question remains of what use there may be for the multimodal, optical-ultrasound nature of the *Dermus SkinScanner* device. Firstly, the combination of optical and ultrasound information has already been shown to increase the diagnostic accuracy of skin tumours.^14^ Similarly, since ultrasound measures of skin inflammation have been shown to correlate well with the semi-quantitatively observed level of skin inflammation,^22,12,41^ even to the extent of detecting subclinical inflammation,^11^ the combined use of optical and ultrasound information may serve to provide better choices for personalized treatment.

In both of the above applications – improved skin cancer diagnosis and skin inflammation scoring – the ability of the *Dermus SkinScanner* to provide registration between the optical and ultrasound images is hypothesized to offer better information fusion. However, since at the current stage of development the optical image quality of the *Dermus SkinScanner* is visibly inferior to those of clinical photos and dermoscopic photos, it would seem advisable to use the optical information of the *Dermus SkinScanner* to register higher-quality optical images to the ultrasound images.

The application of optical-ultrasound image fusion where the *Dermus SkinScanner* arguably has the greatest potential is in preoperative planning. Although the use of OCT (optical coherence tomography) for preoperative planning of nonmelanoma skin cancer is validated by a much larger body of literature,^40^ ultrasound offers a more cost-effective alternative.^42^ The validation of the optical-ultrasound registration methodology of the *Dermus SkinScanner* for treatment planning is the scope of future work, which validation will be helped by a future replacement of the manual scanning mechanism with motorized scanning. Another area of development is providing Doppler imaging capabilities, as this helps assess the inflammatory state of skin and appendages as well as vascularization (see Position Statement 3).^22^ Nevertheless, even at the current stage of development, the optical guidance of the SkinScanner offers the opportunity to find the maximal depth of the lesion (see Position Statement 7)^22^ and images at the same location to ensure reproducibility (see Position Statement 5).^22^

## 5. Conclusion

Preliminary clinical images of a novel optical-ultrasound skin imaging device, the *Dermus SkinScanner*, have been presented. The device is designed to be fully portable, cost-effective and wireless, with the aim of convenient everyday use. Recordings from a variety of common skin lesions show detectability to be comparable to an ultrasound device of similar portability, with both showing image features consistent with those reported in the literature. Since the *Dermus SkinScanner* records both optical and ultrasound images that are registered with each other, this capability presents a number of potential applications, beyond the scope of the current work, including more accurate skin cancer diagnosis, skin inflammation detection and staging, and perhaps more importantly, treatment planning.

Future work is warranted on two fronts. Firstly, the device needs further development for improvement of optical image quality, ultrasound position accuracy using motorization, and artefact reduction using judicious placement of the membrane. The addition of Doppler capability would also help provide additional information on inflammation and vasculature. Secondly, the potential applications of the multimodal optical-ultrasound capability should be studied even at this stage of the device development, including the optical guidance of the ultrasound imaging for reproducible imaging and for finding the maximal depth of the skin cancer tumour. Currently, one of the primary aims of the development is the validation of preoperative skin cancer planning using the device.

In conclusion, the first clinical results from the *Dermus SkinScanner* device presented in this current study warrant further development and investigation of the capabilities of such optical-ultrasound multimodal imaging.

## Data Availability

Data are available on request from the authors.

## Acknowledgements

The authors would like to thank their colleagues at Dermus Kft. for development and documentation of the device, in particular, Eszter Ágoston, György Margitfalvy, Gergely Szikszay-Molnár. The collaboration with Jedlik Innováció Kft under the GINOP-2.1.7-15-2016-02201 programme is gratefully acknowledged.

